# Improving HIV pre-exposure prophylaxis (PrEP) adherence and retention in care: Recommendation development from a national PrEP programme

**DOI:** 10.1101/2022.08.09.22278509

**Authors:** Jennifer MacDonald, Claudia S Estcourt, Paul Flowers, Rak Nandwani, Jamie Frankis, Ingrid Young, Dan Clutterbuck, Jenny Dalrymple, Lisa McDaid, Nicola Steedman, John Saunders

**Author notes:** Corresponding author: Claudia Estcourt, Glasgow Caledonian University, Cowcaddens, Glasgow, G4 0BA, UK. E-mail addresses of authors: JM PF RN JS JF IY DC JD LM NS CE.

## Abstract

**Introduction:** HIV pre-exposure prophylaxis (PrEP) is a key component of HIV combination prevention. Effective prevention needs people to adhere to PrEP during periods of risk and remain in care. However, relevant models of care are under-researched. Using data from the first two years of Scotland’s PrEP programme, we explored barriers and facilitators to PrEP adherence and retention in care and systematically developed evidence-based, theoretically-informed recommendations to enhance future adherence and retention.

**Methods:** We conducted semi-structured interviews and focus groups (09/2018-07/2019) with geographically and demographically diverse patients who were either using/declined/stopped or had been assessed as ineligible for PrEP (n=39), healthcare professionals involved in PrEP provision (n= 54), non-governmental organisation service users (n=9) and staff (n=15) across Scotland. We used thematic analysis to map key barriers and facilitators to priority areas that could enhance adherence and retention in care. Next, we used analytic tools from implementation science (Theoretical Domains Framework, Intervention Functions, Behaviour Change Technique Taxonomy, APEASE criteria) and expert opinion to systematically generate recommendations to enhance future PrEP adherence and retention in care.

**Results:** Barriers and facilitators to adherence and retention in care were diverse and multi-layered. Barriers included perceived complexity of event-based dosing, the tendency for users to stop PrEP before seeking professional support, troublesome side-effects, limited flexibility in the settings, timings, and nature of appointments for follow up, enduring PrEP-related stigma and emerging stigmas around not using PrEP. Facilitators included flexible appointment scheduling, reminders, and processes to follow up non-attenders. We generated 25 wide-ranging but specific recommendations for key stakeholders, for example, emphasising the benefits of PrEP reviews and providing appointments flexibly within individualised PrEP care; using clinic systems to remind/recall PrEP users for review; supporting PrEP conversations among sexual partners; clear guidance on event-based dosing; encouraging/commitment to good PrEP citizenship; and detailed discussion on managing side-effects and care/coping planning activities.

**Conclusions:** PrEP adherence and retention in care is challenging for many people. Such challenges reduce the benefits of PrEP at individual and population levels. Our findings identify and provide solutions to where and how collaborative interventions across public health, clinical, and community practice could address these challenges.

## Introduction

Oral HIV pre-exposure prophylaxis (PrEP, tenofovir disoproxil/emtricitabine) is a highly effective biomedical intervention to reduce HIV acquisition [1] and is central to elimination of HIV transmission. Global implementation of PrEP is accelerating but coverage remains patchy [2]. Evidence to date suggests that adherence to PrEP and retention in care is challenging [1, 3-6]. A systematic review of PrEP trials clearly demonstrated that efficacy is associated with adherence [1]. Other studies show that up to 50% of people who initiate PrEP stop taking it within one year and cessation is associated with younger age, being a transgender women, socio-economic deprivation, lower educational attainment, and drug misuse [7-9]. Cessation of PrEP may happen because of a perceived reduction in HIV acquisition risk, which may or may not be accurate. However, it is unclear how best to identify and support individuals who stop PrEP but remain at, or return to, a risk of HIV acquisition. We need to establish how to encourage adherence to PrEP and retention in care for individuals with ongoing need, and to establish mechanisms through which users can restart PrEP as required.

Learning from large-scale PrEP implementation studies has been limited to date, particularly regarding how services have achieved greatest impact or what could be done to optimise future provision. This is a missed opportunity as real-world studies could be particularly informative more of the PrEP care cascade. To date, attempts to conceptualise the implementation of PrEP have tended to be broad and descriptive, typically categorising the whole of PrEP care into four or five steps within a continuous linear ‘care cascade’ [10-13] or PrEP care pathway. No studies have used conceptualisations of the PrEP care cascade as the starting point for systematic and focussed service improvement.

Scotland became one of the first countries worldwide to implement a national PrEP programme [14]. At the time, there were around 4600 people living with HIV attending specialist care in Scotland [15] and 228 people newly diagnosed with HIV each year, half of whom were gay, bisexual, and other men who have sex with men (GBMSM) [16]. From July 2017, PrEP and all associated monitoring were made available as part of broader HIV combination prevention and sexual health care, free at point of access almost exclusively through sexual health clinics, to those at greatest risk of HIV acquisition [17]. Prescribing followed specialist association guidance [18], but services developed their own local models of delivery, largely within existing budgets. These broadly involved: (1) identifying a patient as a PrEP candidate; (2) provision of PrEP information, baseline screening for HIV, other blood borne viruses (BBVs), sexually transmitted infections (STIs), and renal function; (3) prescribing and dispensing PrEP; and (4) regular in person reviews for HIV, BBV, and STI testing, renal monitoring, adherence support, wider sexual health promotion, and PrEP prescribing [18]. Quantitative outcomes from the Programme have been reported as part of routine surveillance [17,19,20] and within a detailed epidemiological study [21].

We conducted an evaluation of the first two years of Scotland’s PrEP programme. Our approach divided the PrEP care cascade into three sections; awareness and access, initiation and uptake (both described elsewhere) and adherence and retention in care. Here we focussed on adherence and retention in care as a broad domain potentially in need of behaviour change interventions to enhance implementation. We defined *adherence* as taking PrEP in line with medical advice / using PrEP appropriately (critical for efficacy) and *retention in care* as attending PrEP review appointments and staying on PrEP during periods of risk.

We addressed the following research questions:

1. Within PrEP care pathways, where should we intervene (priority areas) to improve PrEP adherence and retention in care?
2. What are the barriers and facilitators to implementing the priority areas for PrEP adherence and retention in care?
3. Which evidence-based and theoretically informed recommendations could improve PrEP adherence and retention in care?

## Methods

This study involved Stage 1: a retrospective qualitative process evaluation within a larger natural experimental design study evaluating PrEP implementation in Scotland (research questions 1 and 2), and Stage 2: development of recommendations to improve PrEP adherence and retention in care, using systematic intervention development approaches (research question 3).

## Data collection

### Participants

We used multi-perspective purposive sampling to understand the implementation of PrEP adherence and retention in care from diverse viewpoints. In total, 117 participants took part in individual semi-structured telephone interviews (n=71) or in one of 10 group discussions (n=46) (September 2018-July 2019). The sample comprised: 39 patients; 54 healthcare professionals; nine non-governmental organisation (NGO) service users; and 15 NGO staff from across Scotland. All NGOs had an HIV prevention remit and served GBMSM, trans, and/or Black African communities. Group discussions included one type of stakeholder only.

Patients were either using PrEP (n=23, 59%) or had declined (n=5, 13%), stopped (n=6, 15%), or been assessed as ineligible (n=5, 13%) for PrEP. PrEP users included those who took PrEP daily, event-based or both ways. They ranged in age from 20-72 years with just over half (n=21, 54%) between 25-34 years. All self-identified as gay or bisexual men, the majority of whom (n=34, 87%) were cisgender. Almost all were of ‘White British’ (n=31, 80%) or ‘Other White’ (n=7, 18%) ethnicity. Two thirds reported a university degree as their highest level of education (n=26, 67%) and the majority were in employment (n=34, 87%). The patient areas of residence reflected a mix of relative affluence and deprivation although the most (n=5, 16.7%) and least (n=3, 10%) deprived quintiles (according to Scottish Index of Multiple Deprivation (SIMD), which divides areas into five subgroups according to the extent to which an area is “deprived” [22]) were under-represented and patients predominantly resided in the middle three quintiles (73%) (data missing for 9 participants). Healthcare professionals were all involved in PrEP implementation in a mix of rural (n=12, 22%), semi-rural/urban (n=8, 15%), or urban (n=34, 63%) settings, largely reflecting the wider Scottish population distribution. They included specialist sexual health doctors and nurses of various grades, some with national PrEP roles, PrEP prescribing general practitioners (who prescribed PrEP where there was no sexual health service on their Scottish island), health promotion officers, a midwife, and a clinical secretary responsible for PrEP-related administration. NGO service users were all of Black African ethnicity, predominantly cis-gender women, and not using PrEP.

### Recruitment

Healthcare professionals offered patients the opportunity to take part in the study during routine consultations taking place in four of the 14 regional health boards (responsible for the protection and improvement of their population’s health)) providing over 90% of PrEP related care in Scotland. NGO service users who were either engaged with NGOs *and* attending sexual health clinics (classed as patients above) or only engaged with NGO services (classed as NGO service users above) were invited to participate via interactions with NGO staff. We recruited these and other NGO staff and healthcare professionals across all of Scotland’s 14 regional health boards by email invitation.

### Procedure

All participants provided informed verbal or written consent immediately prior to the interviews /group discussions. We collected data with the aid of a topic guide that included open-ended questions designed to explore participants’ experiences and perceptions of PrEP adherence and retention in care, rather than questions based on any theoretical concepts anticipated to influence implementation. Where possible within the group discussions, dialogue between participants was encouraged rather than between facilitators and participants. All participants talked from their own and others’ perspectives; data were taken at face value. Patients were offered a £30 shopping voucher as reimbursement for their time.

Data collection was led by JM, with input from experienced qualitative researchers, PF, IY, and JF. JM, PF, IY, and JF reviewed and discussed early transcripts for quality assurance purposes. All interviews and group discussions were audio recorded, transcribed verbatim, anonymised, and imported into NVivo software for analysis.

## Data analysis

### Stage 1

#### 1. Within PrEP care pathways, where should we intervene (priority areas) to improve PrEP adherence and retention in care?

Firstly, we used the Action, Actor, Context, Target, Time framework [23], to conceptualise the sequential actors, actions, settings, and processes that constituted PrEP adherence and retention in care. Secondly, we iteratively created a series of visualisations of the overall behavioural system of PrEP adherence and retention in care using available UK guidance on best clinical practice in PrEP provision [18] and transcripts of early interviews and group discussions. Thirdly, we comprehensively assessed the breadth and depth of data relating to the patient pathway through PrEP adherence and retention in care. Fourthly, we (PF & JM) ranked the most important areas which were considered to be amenable to change to create priority areas for intervention This stage combined the earlier findings with input from the specialist doctor team members who had real-world clinical experience of providing PrEP services in assorted settings (CSE, RN, JS). This stage ended with the identification of nine priority areas for recommendation development.

#### 2. What are the barriers and facilitators to implementing the priority areas for PrEP adherence and retention in care?

We (JM and PF) conducted deductive thematic analysis [24] of the qualitative data concerning barriers and facilitators for each priority area. We used the relative frequency of barriers and facilitators to manage the volume of findings and to ensure we focussed only on those that were deemed most important. This stage ended with the identification of the major barriers and facilitators for priority areas relating to adherence and retention in care.

### Stage 2

#### 3. Which evidence-based and theoretically informed recommendations could improve PrEP adherence and retention in care?

We treated each of the priority areas independently and analysed each one separately. Firstly, we entered the key barriers and facilitators into a matrix. Secondly, we used the Behaviour Change Wheel (BCW) approach [25], to characterise behaviour change components of PrEP care and systematically coded the key barriers and facilitators for each priority area. Thirdly, we used the Theoretical Domains Framework (TDF) [26] to theorise the key barriers and facilitators. Fourthly, we specified corresponding Intervention Functions (broad ways of intervening relevant to the theoretical domains) and used the Behaviour Change Technique (BCT) and corresponding Taxonomy (BCTT) [27] to describe, in detail and using a standardised language, potential intervention content that may be helpful to operationalise the Intervention Functions, address key barriers and facilitators, and enhance implementation. This created an initial “long-list” of recommendations. All coding and drafting of recommendations were completed by JM and double-checked for accuracy, validity, and credibility by PF. Any disagreements were discussed until consensus was reached.

Finally, clinical expert team members (CE, RN, JS) scrutinised, sense-checked, and shortlisted the long list of initial recommendations using the APEASE criteria [28]. This resulted in the introduction of some new recommendations, in addition to minor amendments to or merging/deleting of existing recommendations.

## Ethical considerations

The Glasgow Caledonian University Research Ethics Committee (HLS/NCH/17/037, HLS/NCH/17/038, HLS/NCH/17/044) and the South East Scotland National Health Service Research Ethics Committee (18/SS/0075, R&D GN18HS368) provided ethical approval.

## Results

### 1. Within PrEP care pathways, where should we intervene (priority areas) to improve PrEP adherence and retention in care?

We identified 10 priority areas for intervention within the final visualised behavioural system (Table 1, Figure 1 & Appendix 1) of a typical PrEP care pathway for adherence (n=2) and retention in care (n=8). These priority areas involved two actors (PrEP providers and PrEP users). Six were interactional (1, 4, 5, 6, 8, and 9) and concerned supporting effective PrEP use, assessing ongoing eligibility for PrEP, discussing and addressing wider sexual health issues, communicating the decision to not provide further PrEP, and exploring reasons for wanting to stop/stopping PrEP. Four were more individually oriented (2, 3, 7, and 10) and concerned PrEP users taking PrEP in line with medical advice, attending PrEP reviews, continuing to use PrEP for as long as required, and stopping PrEP safely.

**Table 1.**
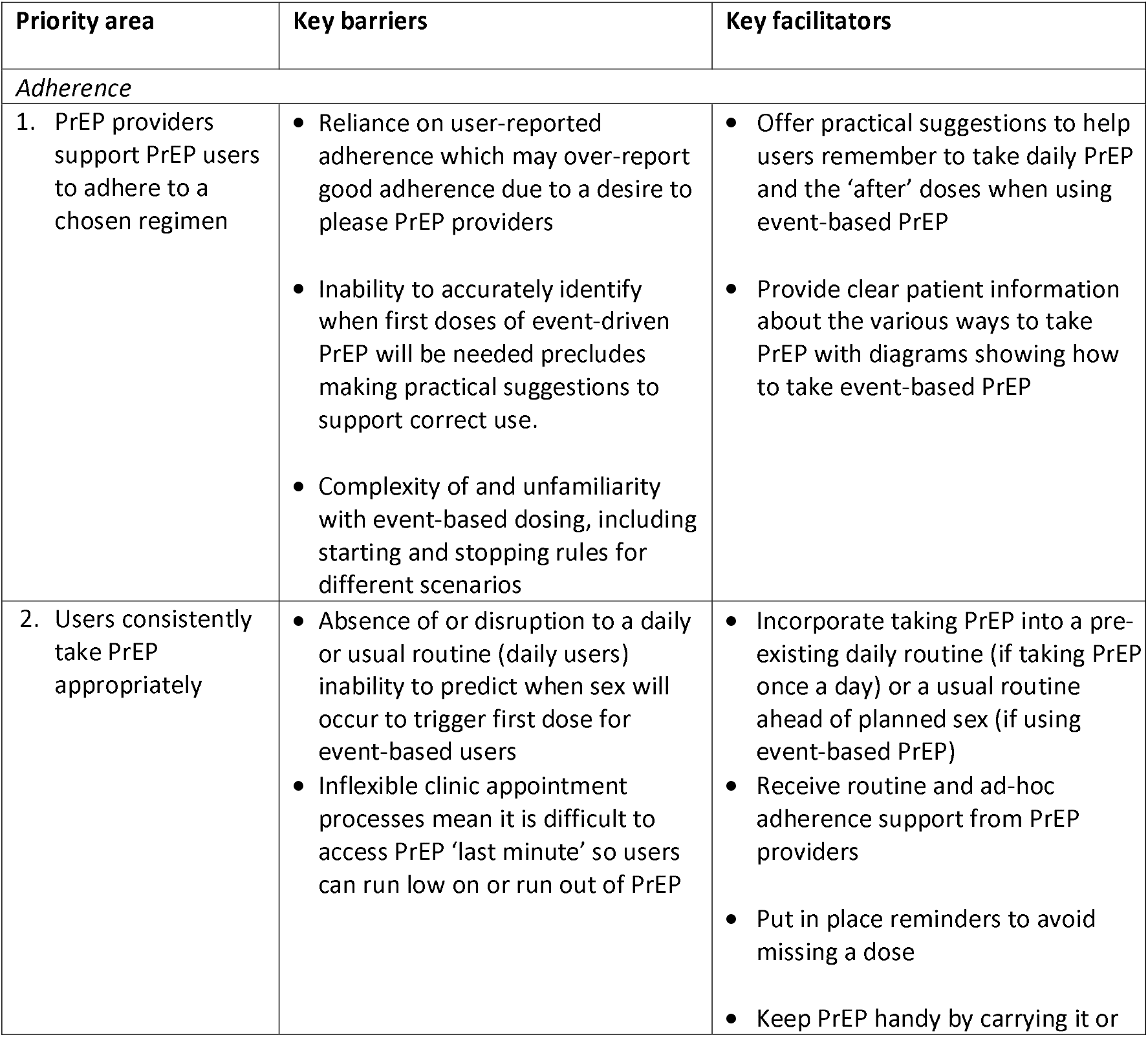
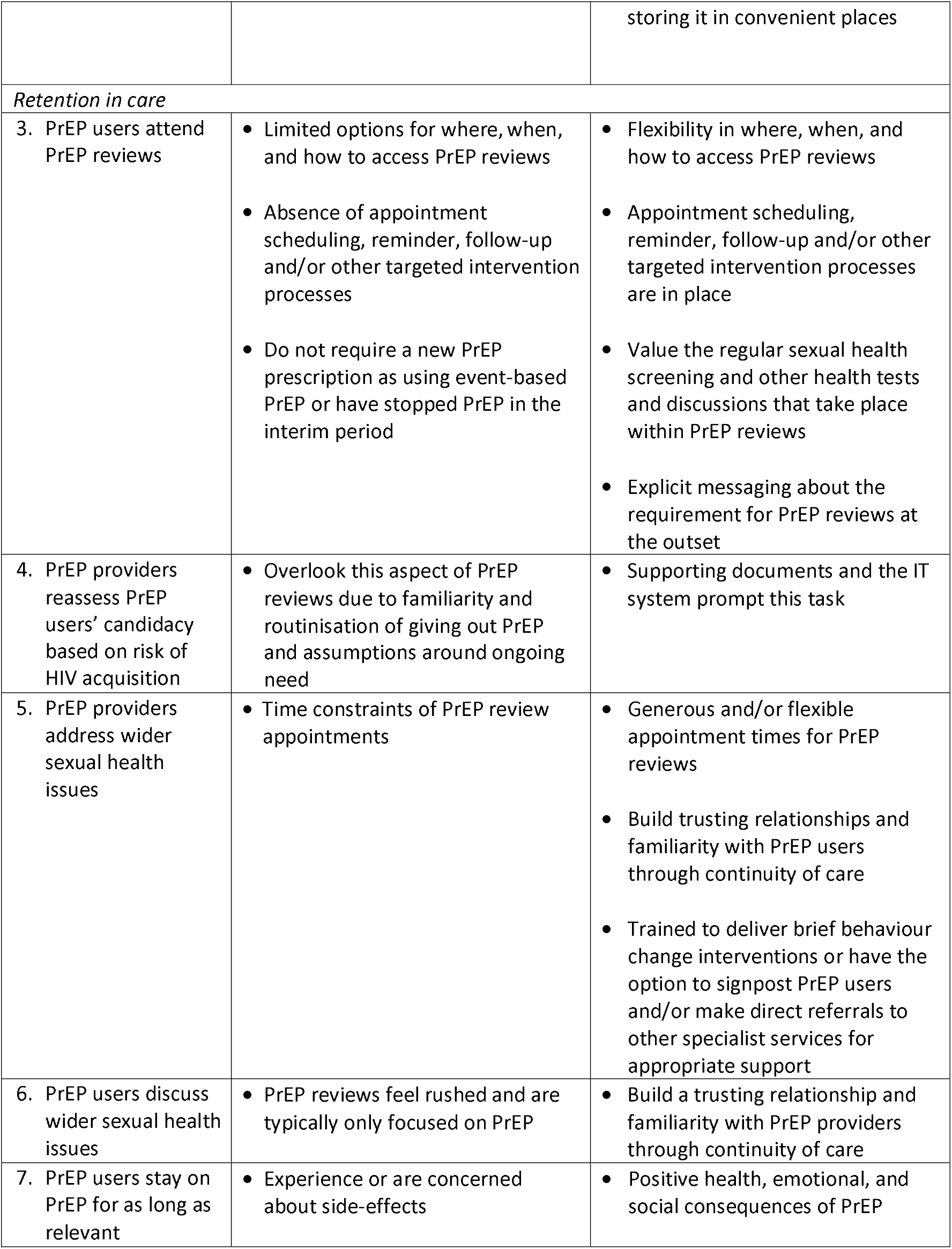
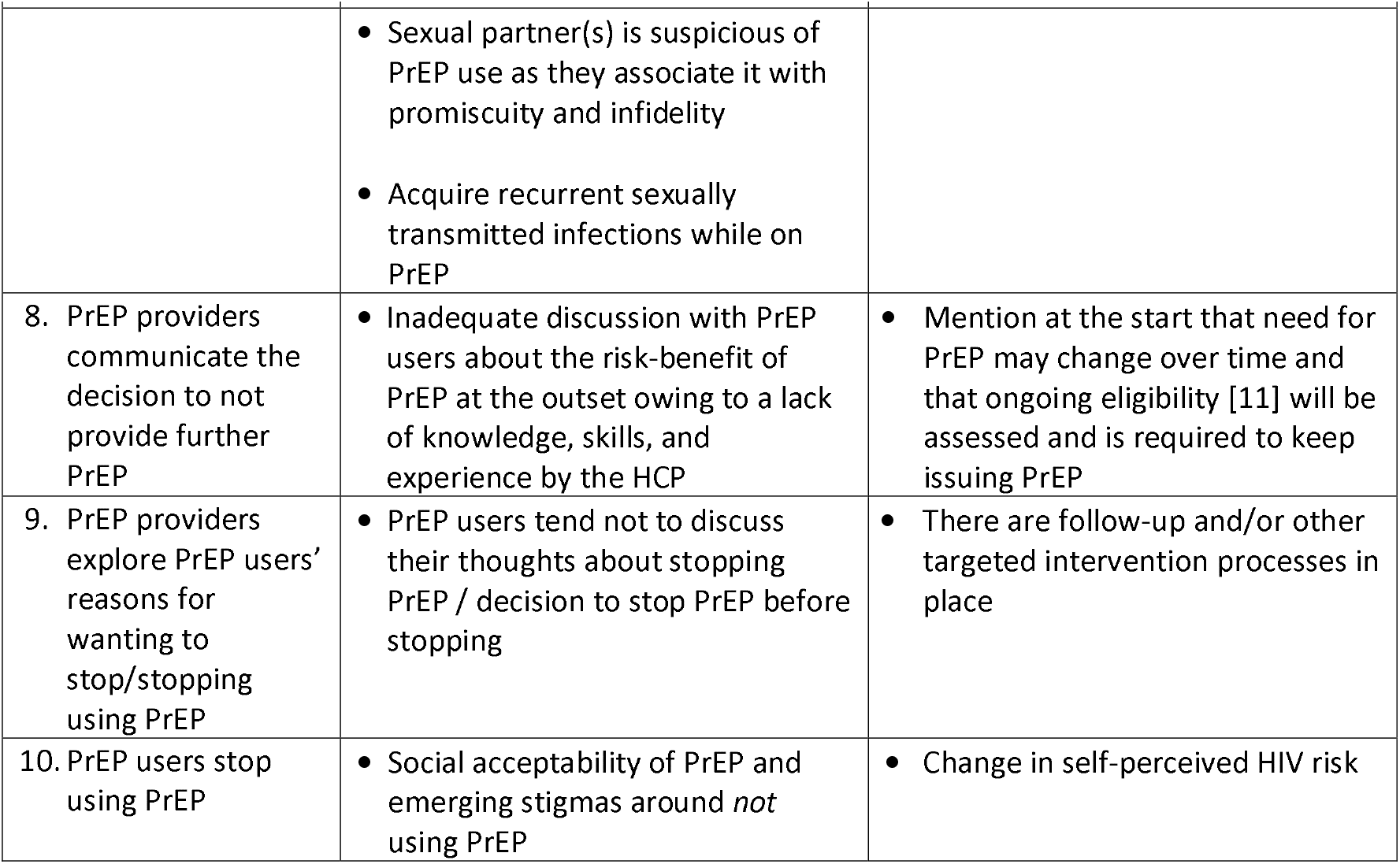
**Key barriers and facilitators to the priority areas for PrEP adherence and retention in care**.

**Figure 1.**
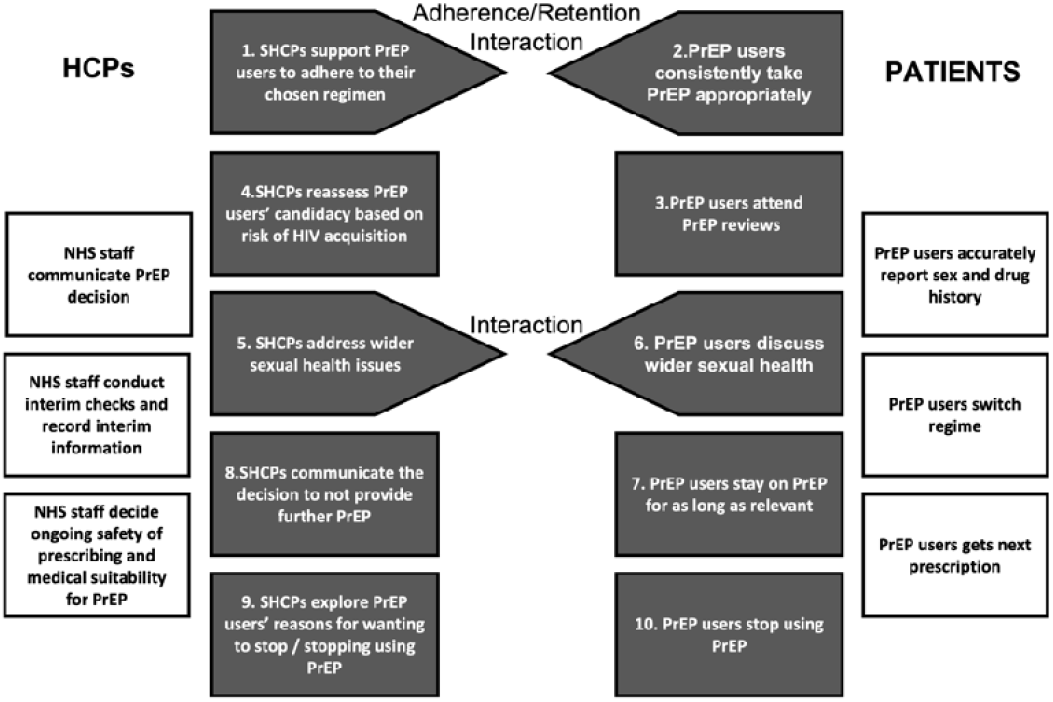
A schematic of the behavioural system of adherence and retention in care. White boxes – not selected as a priority area as not considered amenable to change Black boxes – selected as a priority area Arrowed Boxes – demonstrate priority areas that interact

### 2. What are the barriers and facilitators to implementing the priority areas for PrEP adherence and retention in care?

The key barriers and facilitators relating to our priority areas, which were multi-levelled and ranged from the macro to the micro, are shown in Table 1.

### 3. Which evidence-based and theoretically informed recommendations could improve PrEP adherence and retention in care?

We generated an initial 51 recommendations to address the priority areas identified (see Appendix 2 for the full evidence table of key barriers and facilitators to priority areas, TDF domains, Intervention Functions, BCTs, and initial recommendations) which we reduced to 25 final recommendations after applying the APEASE criteria (Table 2).

**Table 2.**
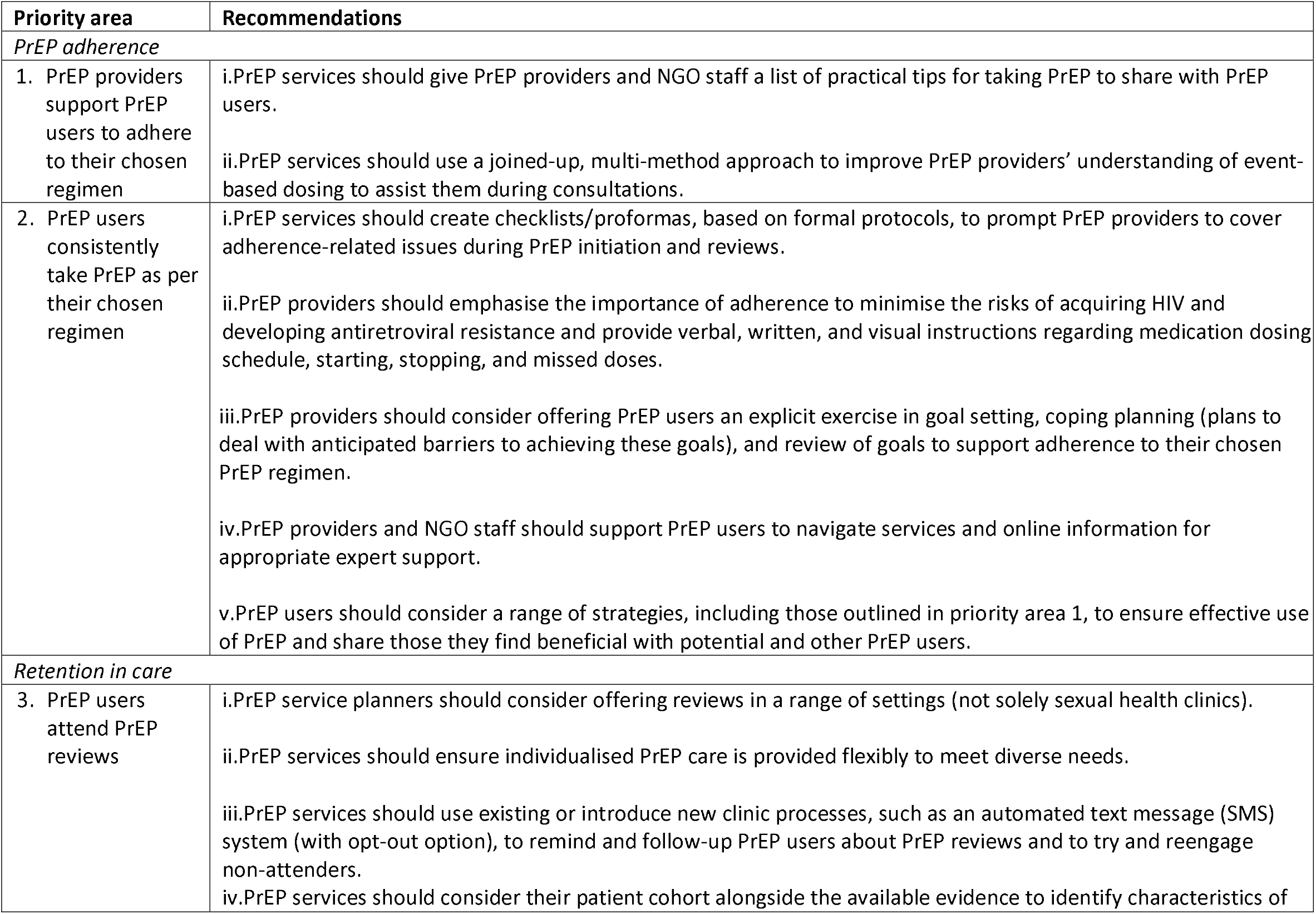
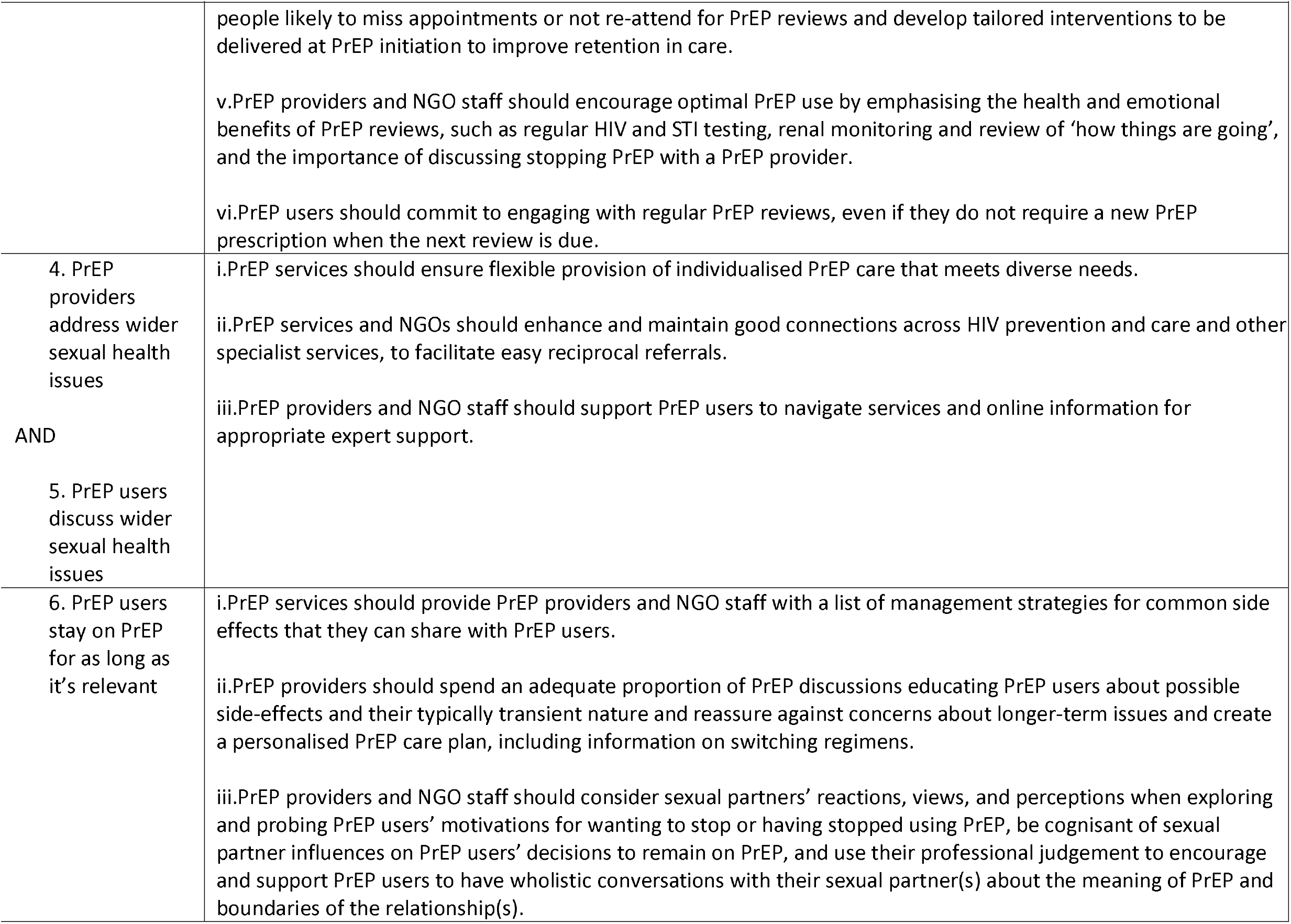
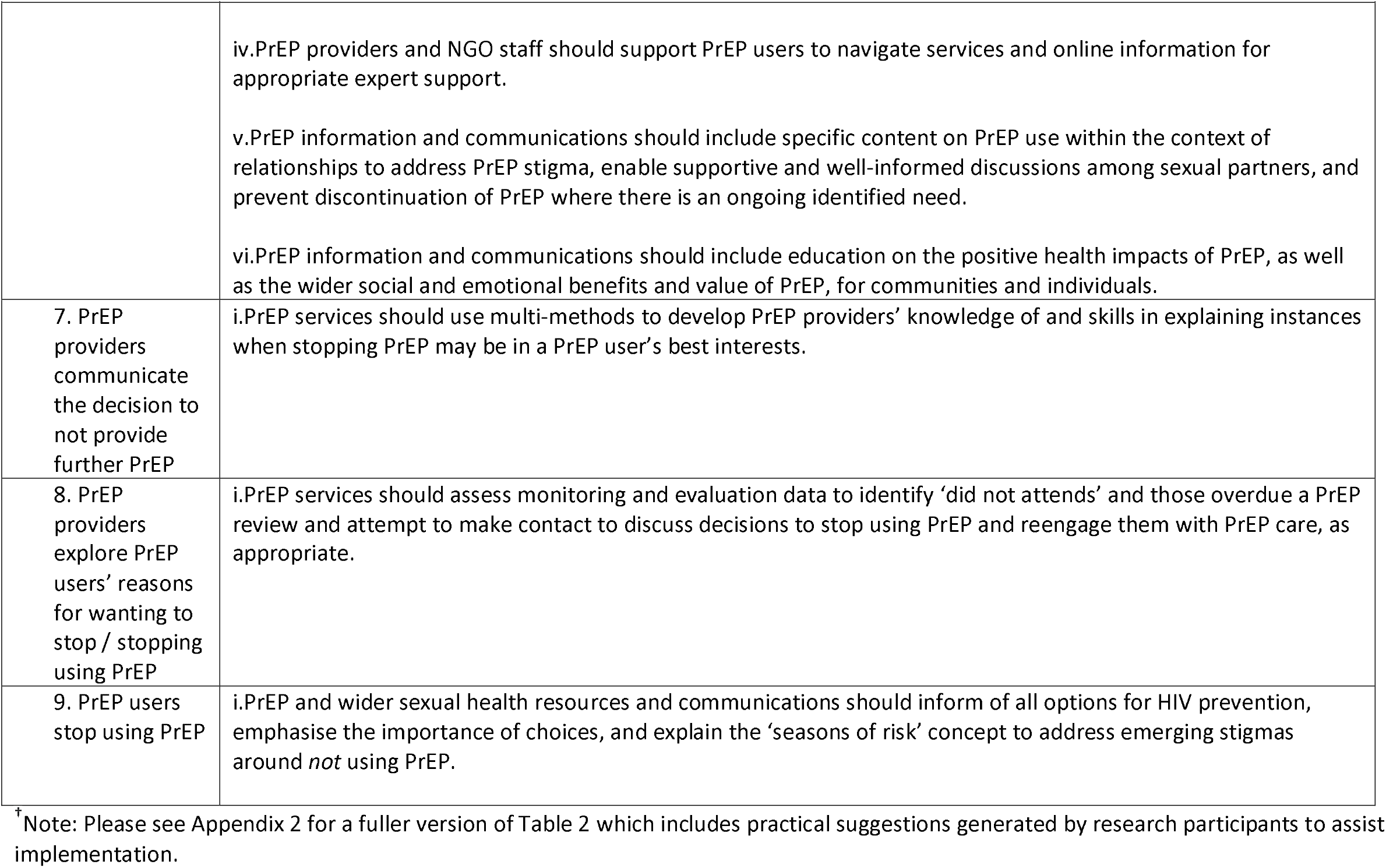
**Final evidence-based and theoretically informed recommendations to improve PrEP adherence and retention in care**^†^

| Priority area | Recommendations |
| --- | --- |
| *PrEP adherence* |  |
| 1. PrEP providers support PrEP users to adhere to their chosen regimen | i.PrEP services should give PrEP providers and NGO staff a list of practical tips for taking PrEP to share with PrEP users.<br>ii.PrEP services should use a joined-up, multi-method approach to improve PrEP providers' understanding of event-based dosing to assist them during consultations. |
| 2. PrEP users consistently take PrEP as per their chosen regimen | i.PrEP services should create checklists/proformas, based on formal protocols, to prompt PrEP providers to cover adherence-related issues during PrEP initiation and reviews.<br>ii.PrEP providers should emphasise the importance of adherence to minimise the risks of acquiring HIV and developing antiretroviral resistance and provide verbal, written, and visual instructions regarding medication dosing schedule, starting, stopping, and missed doses.<br>iii.PrEP providers should consider offering PrEP users an explicit exercise in goal setting, coping planning (plans to deal with anticipated barriers to achieving these goals), and review of goals to support adherence to their chosen PrEP regimen.<br>iv.PrEP providers and NGO staff should support PrEP users to navigate services and online information for appropriate expert support.<br>v.PrEP users should consider a range of strategies, including those outlined in priority area 1, to ensure effective use of PrEP and share those they find beneficial with potential and other PrEP users. |
| *Retention in care* |  |
| 3. PrEP users attend PrEP reviews | i.PrEP service planners should consider offering reviews in a range of settings (not solely sexual health clinics).<br>ii.PrEP services should ensure individualised PrEP care is provided flexibly to meet diverse needs.<br>iii.PrEP services should use existing or introduce new clinic processes, such as an automated text message (SMS) system (with opt-out option), to remind and follow-up PrEP users about PrEP reviews and to try and reengage non-attenders.<br>iv.PrEP services should consider their patient cohort alongside the available evidence to identify characteristics of |
|  | people likely to miss appointments or not re-attend for PrEP reviews and develop tailored interventions to be delivered at PrEP initiation to improve retention in care.<br>v.PrEP providers and NGO staff should encourage optimal PrEP use by emphasising the health and emotional benefits of PrEP reviews, such as regular HIV and STI testing, renal monitoring and review of ‘how things are going’, and the importance of discussing stopping PrEP with a PrEP provider.<br>vi.PrEP users should commit to engaging with regular PrEP reviews, even if they do not require a new PrEP prescription when the next review is due. |
| 4. PrEP providers address wider sexual health issues<br>AND<br>5. PrEP users discuss wider sexual health issues | i.PrEP services should ensure flexible provision of individualised PrEP care that meets diverse needs.<br>ii.PrEP services and NGOs should enhance and maintain good connections across HIV prevention and care and other specialist services, to facilitate easy reciprocal referrals.<br>iii.PrEP providers and NGO staff should support PrEP users to navigate services and online information for appropriate expert support. |
| 6. PrEP users stay on PrEP for as long as it’s relevant | i.PrEP services should provide PrEP providers and NGO staff with a list of management strategies for common side effects that they can share with PrEP users.<br>ii.PrEP providers should spend an adequate proportion of PrEP discussions educating PrEP users about possible side-effects and their typically transient nature and reassure against concerns about longer-term issues and create a personalised PrEP care plan, including information on switching regimens.<br>iii.PrEP providers and NGO staff should consider sexual partners’ reactions, views, and perceptions when exploring and probing PrEP users’ motivations for wanting to stop or having stopped using PrEP, be cognisant of sexual partner influences on PrEP users’ decisions to remain on PrEP, and use their professional judgement to encourage and support PrEP users to have wholistic conversations with their sexual partner(s) about the meaning of PrEP and boundaries of the relationship(s). |
|  | iv. PrEP providers and NGO staff should support PrEP users to navigate services and online information for appropriate expert support.<br>v. PrEP information and communications should include specific content on PrEP use within the context of relationships to address PrEP stigma, enable supportive and well-informed discussions among sexual partners, and prevent discontinuation of PrEP where there is an ongoing identified need.<br>vi. PrEP information and communications should include education on the positive health impacts of PrEP, as well as the wider social and emotional benefits and value of PrEP, for communities and individuals. |
| 7. PrEP providers communicate the decision to not provide further PrEP | i. PrEP services should use multi-methods to develop PrEP providers' knowledge of and skills in explaining instances when stopping PrEP may be in a PrEP user's best interests. |
| 8. PrEP providers explore PrEP users' reasons for wanting to stop / stopping using PrEP | i. PrEP services should assess monitoring and evaluation data to identify 'did not attend' and those overdue a PrEP review and attempt to make contact to discuss decisions to stop using PrEP and reengage them with PrEP care, as appropriate. |
| 9. PrEP users stop using PrEP | i. PrEP and wider sexual health resources and communications should inform of all options for HIV prevention, emphasise the importance of choices, and explain the 'seasons of risk' concept to address emerging stigmas around *not* using PrEP. |

† Note: Please see Appendix 2 for a fuller version of Table 2 which includes practical suggestions generated by research participants to assist implementation.

No recommendations for priority area four (PrEP providers reassess PrEP users’ candidacy for PrEP based on risk of HIV acquisition) were retained because it is a required element of care.

## Discussion

### Main findings

We identified nine priority areas in the PrEP care cascade which could be optimised to improve adherence and retention in care. PrEP users, health care professionals and NGO staff and clients identified multiple barriers and facilitators to effective engagement with these priority areas. Using robust methodology with tools from implementation science, we derived 25 specific recommendations to enhance future PrEP implementation. Recommendations range from those at the “micro-level” within interactions between health care professionals and PrEP users, which broadly encompassed tailoring PrEP care to the individual, to higher level suggestions for collaboration across agencies and provision of a PrEP in a variety of different settings to meet diverse needs.

### Strengths and weaknesses

This large study involved a wide range of clinical and non-clinical stakeholders with varied perspectives and priorities, within a national PrEP programme. We focussed on adherence to PrEP and retention in care which can be problematic steps within the PrEP care cascade at which there are known to be inequity in outcomes for key vulnerable populations [6]. Our innovative approach draws directly on staff and patient perspectives and uses the cumulative knowledge embodied within theories of implementation [29] and contributes to implementation science through the shared language and depiction of core concepts (i.e., intervention functions and behaviour change techniques).

We acknowledge that data were generated from a single country in which PrEP provision was provided free of charge within sexual health clinics. However, many of the recommendations, such as those which relate to tailoring PrEP support to the individual, flexible appointments and information are likely to be applicable in most settings in which PrEP is provided, even when PrEP is funded by the individual. In contrast, recommendations which specifically relate to sexual health clinic-based PrEP delivery, may lack wider applicability. We conducted the study in first two years of the PrEP programme and so findings reflect early stage implementation. Some barriers and facilitators may change as the programme matures, for example as users and providers become more familiar with event-based dosing. The participants using PrEP were largely representative of people on PrEP in Scotland at the time and, despite our efforts, women, trans and gender diverse people are relatively underrepresented. This lack of sample diversity means that the experience and perspectives of health care professionals may largely only relate to providing PrEP care to cisgender gay, bisexual and other men who have sex with men.

### Findings in context of other studies

Our findings build on those from several other studies which have highlighted various barriers to PrEP adherence and retention in care and our findings are in keeping with many of these [4,30-31], which provides legitimacy to our findings. Furthermore, our recommendations are broadly aligned with elements of recommendations from other authors and public health agencies, (for example, co-production of materials [32] and support in navigating health systems, e.g., Prepster [33]). Similarly, embedding PrEP delivery within combination prevention together with a focus on broader sexual wellbeing was successful in maintaining young men who have sex with men of colour on PrEP in a small feasibility pilot [34]. It is also a model of care recommended within PrEP guidelines [12,35] and is in keeping with several of our recommendations. The use of text reminders to attend healthcare appointments and adhere to medication has been successfully used in many health areas, including for PrEP, supporting our recommendation to use automated text reminders [36,37]. Some promising interventions have not been deployed in Scotland hence do not have recommendations for example, the use of peer navigators to assist people to engage with PrEP which was found useful for some [38]. To our knowledge, none of the previously published guidance has used the rigorous approach to generating recommendations that we took [39,40] or provided such a comprehensive list of recommendations focussed on this stage of the PrEP care cascade.

There are examples of effective interventions to improve medication adherence for other disease areas including for people living with HIV taking antiretroviral medication and other conditions requiring long term drug therapy [41-43]. Although these relate to people already diagnosed with a chronic condition which requires long term medication rather than people trying to avoid an infection, there are similarities with our findings. Adaptation of these existing interventions could be useful to improve PrEP adherence and retention in care [44] and vice versa. However, a Cochrane review of improving adherence to and continuation of hormonal contraception, which might better approximate PrEP as it relates to prevention rather than treatment, provided less overlap in findings. For example, intensive counselling and reminders may result in only a slight increase in continuation of hormonal contraception although the effect varied by contraception method [45]. However, to date, interventional studies based on published recommendations, and designed to overcome barriers to improve PrEP adherence and retention specifically, are lacking and robust evaluation of the impact of these approaches is scarce.

### Implications for policy and practice

Many of our recommendations highlight the importance of supporting the individual and understanding their concerns and priorities, together with tailored advice and activities to enhance their understanding of PrEP with discussion of specific strategies to help with ensuring that PrEP is taken appropriately and safely at times of risk, through adherence to suitable dosing regimen(s). All of these are in keeping with a person-centred approach to care. However, we acknowledge that these activities take time within consultations and services may lack adequate resources to fully provide this as they are currently organised. Within the UK context, sexual health service delivery has changed significantly during the SAR-CoV2 pandemic with face-to-face appointments being reserved for people who are symptomatic and or have more complex needs. PrEP services have largely shifted to telephone models [46]. The opportunity to deliver some of our recommendations may be more challenging should services continue with more remote and light-touch models of care but are no less important. However, this could be an opportunity to commission services through community-based organisations, such as the use of peer navigators. Although the future provision of long-acting PrEP formulations could reduce adherence demands in some respects, there will still be a need for regular monitoring and adherence support. Detailed recommendations to enhance adherence such as these may be even more needed.

Across PrEP services more broadly, health care professionals and NGO staff may benefit from training to improve their skills and could usefully learn from each other. NGO staff could play a key role in cultural competency training as well as helping to extend the reach of PrEP to key populations that could benefit, thereby helping to reduce inequalities in provision. In settings where generic medication is available, the costs of providing this support may outstrip drug costs and would need to be appropriately funded in the health care and NGO setting.

## Conclusions

The potential for PrEP to have a major impact on HIV transmission relies on people adhering to it and remaining in active follow up as appropriate to their needs. These recommendations could directly enhance the quality of PrEP care at an individual patient level and inform development of interventions to improve adherence and retention in care at programme-level. More work is needed with people from a wide range of groups who could benefit from PrEP (women, trans and non-binary communities, people who inject drugs, migrant communities.) to ensure that recommendations and interventions are appropriate to all key groups and to avoid inadvertently widening existing health inequalities. Future work should include robust evaluation of implemented recommendations.

## Data Availability

Data that supports the findings and suggestions of this study are explained within the the supplementary material for this article, which presents elements of the original dataset.

## Authors’ contributions

All authors contributed to the conception and design of the studies, interpretation of findings, revision of the manuscript and approved the final version. Specific additional contributions are as follows and marked where appropriate in the manuscript: CSE was principal investigator and involved in all stages of the research and wrote the initial draft of the manuscript. JM led the study day to day and undertook all research activities including data collection and analysis under the supervision of PF and CSE. JS, RN, DC, NS and CSE provided expert clinical interpretation. IY and JF contributed to data collection and analysis. JD and JF led the ethical approval application.

## Author information [Optional]

Claudia Estcourt is monitoring and research lead for Scotland’s national PrEP Programme and is part of Scotland’s HIV transmission elimination oversight group. She was Programme Steering Committee Chair for England’s Impact Trial. She is co-author of BHIVA/BASHH guidelines on the use of HIV pre-exposure prophylaxis (PrEP), 2022 and led European Centre for Disease Prevention and control (ECDC) HIV Pre-Exposure Prophylaxis in the EU/EEA and the UK: implementation, standards and monitoring, technical guidance, 2021.

Rak Nandwani chaired the Scotland PrEP Short Life Working Group in 2016. He currently chairs the HIV transmission elimination oversight group which will submit proposals to Scottish Government in 2022. He is also a non-executive director of the Board of Public Health Scotland. Ingrid Young was on the Scotland PrEP Short Life Working group in 2016, was a co-author of the 2018 and forthcoming (2022) BHIVA-BASHH guidelines on the use of Pre-exposure prophylaxis (PrEP).

Nicola Steedman was on the Scotland PrEP Short Life Working Group in 2016 (as Senior Medical Officer for Scottish Government). She co-Chaired the Scottish National PrEP Monitoring and Research Group (with Professor Estcourt) and is currently Deputy Chief Medical Officer for Scottish Government with a remit which includes Sexual Health and Bloodborne Viruses.

## Acknowledgements

We are very grateful to the users, patients and staff of sexual health services in all 14 Health Boards, Drs Ruth Holman, Dan Clutterbuck, Maggie Gurney, Nil Banerjee, Pauline McGough, Daniela Brawley, Kirsty Abu-Rajab, Hame Lata, Anne McLellan, Alison Currie, Sharon Cameron, Hilary MacPherson, Janice Irvine, Graham Leslie, Ciara Cunningham, Maggie Watts. We thank staff and users of HIV Scotland; Waverley Care (SX Project and African Health Project); THT Scotland; Hwupenyu Health and Wellbeing; and Scottish Trans Alliance. We thank Nathan Sparling and Jacqueline Gray for their contributions to the research process.

## Additional files [Optional]

Appendix 1: Full evidence tables of key barriers and facilitators to the priority areas, TDF domains, Intervention Functions, BCTs, and original recommendations.

## List of abbreviations [Optional]

APEASE: Affordability, Practicability, Effectiveness and cost-effectiveness, Acceptability, Side-effects and safety, Equity
BBV: Blood borne viruses
BCT: Behaviour Change Technique
BCTT: Behaviour Change Technique Taxonomy
BCW: Behaviour Change Wheel
GBMSM: Gay, bisexual, and other men who have sex with men
HIV: Human immunodeficiency virus
PrEP: Pre-exposure prophylaxis
STI: Sexually transmitted infection
TDF: Theoretical Domains Framework

