## Appendix 1 for "Improving HIV pre-exposure prophylaxis (PrEP) adherence and retention in care: Recommendation development from a national PrEP programme"

**Appendix 1 - Full evidence tables of key barriers and facilitators to the priority areas, TDF domains, Intervention Functions, BCTs, and original recommendations.**

**Table S1 -Priority area 1- A BCW analysis of ‘PrEP providers support PrEP users to adhere to their chosen regimen’**

| **Barriers** | **Facilitators** | **Indicative quotes** | **TDF domains** | **Intervention Functions** | **Potential BCTs**  **Numbers = BCTs** | **Initial recommendations for those considering implementing PrEP at scale**  **Accept/Reject/Modify**  **Numbers in brackets = BCTs** | **Post APEASE and expert input decision**    **Accept/Reject/Modify** | **Agreed final recommendations** **for those considering implementing PrEP at scale** |
| --- | --- | --- | --- | --- | --- | --- | --- | --- |
| PrEP providers find it difficult to support PrEP users to adhere to their chosen regimen because they have to rely solely on what PrEP users tell them and their reported adherence may differ to their actual adherence due to a desire to please PrEP providers | **--** | “*With PrEP it’s very difficult, you don’t have an objective measure of their adherence so you only know what patients are telling you and generally patients want to please their clinicians so they will say to you, oh, no, I’ve been taking it. So whether that’s accurate or not I don’t know*.” (Sexual healthcare professional) | Social influences | Environmental restructuring  Enablement | 6.2 Social comparison  12.2 Restructure the social environment  1.2 Problem solving  7.1 Prompts/cues | 39. Sexual healthcare professionals should draw attention to the challenges of taking PrEP based on what they know about the experiences of other PrEP users (6.2) and cultivate a no blame and non-judgemental approach to encourage an open dialogue about any adherence issues or concerns (12.2)  40. Sexual healthcare professionals should engage PrEP users in a discussion of factors that (could) influence adherence and generate suitable solutions (1.2) then document the discussion in the electronic patient record as a useful basis for opening the adherence conversation at the next appointment (7.1) | 39. Reject (discussed) – part of routine practice not PrEP specific.  40. Reject (discussed) – not sure that the electronic patient record is a good place for opening conversations about adherence. | Do not take forwards  Do not take forwards |
| PrEP providers find it difficult to support PrEP users to adhere to their chosen regimen because the variability in PrEP users’ circumstances surrounding the initiation of event-based dosing precludes making practical suggestions to trigger the first dose | PrEP providers find it easy to support PrEP users to adhere to their chosen regimen because they can offer practical suggestions to help PrEP users remember to take daily PrEP and the ‘after’ doses, if using event-based PrEP | “*With daily dosing it’s like with the pill, we tell them you can put an alarm on your phone or something or if you’re on another medicine, take it at the same time, you know, just ways that they can remember it. The event-based one is trickier, you know, to think of triggers that can help them remember to do it*.” (Sexual healthcare professional) | SRejects  Environmental context and resources | Training  Education  Enablement | 4.4 Behavioural experiments  4.1nstruction on how to perform the behaviour | 41. Sexual healthcare professionals could suggest that people using event-based PrEP test different approaches to trigger their initial dose and note which approach is most successful (4.4)  41. Sexual health services should provide sexual healthcare professionals with a list of practical tips (e.g. in a national patient information booklet, wallet-sized insert) that they can share and discuss with PrEP users to encourage adherence to a daily PrEP regimen or the ‘after’ doses, if using event-based PrEP (4.1) | 41. Keep (discussed) – merge with others marked 41.  41. Keep (discussed) – add a new sentence at the end ‘Practical tips could include advising PrEP users to:’ then list other recommendations marked 41. | PrEP services should provide PrEP providers and NGO staff with a list of practical tips for taking PrEP to share with PrEP users. *Strategies for daily PrEP and the ‘after’ doses of event-based PrEP include: formulating an ‘if-then’ plan that links taking PrEP once a day to a specific task (e.g. brushing teeth) which remains constant even in the absence of or disruption to a daily routine; marking PrEP use on a calendar or recording it in a diary; setting reminder alarms and/or using a pill organiser; and keeping PrEP handy by carrying it and/or storing it in convenient places. A strategy for starting event-based PrEP could be to test different approaches to trigger the initial dose and note which approach is the most successful* |
| PrEP providers find it difficult to support PrEP users to adhere to their chosen regimen because of the complexity of and unfamiliarity with event-based dosing (e.g. when to start, stopping rules for different scenarios) | PrEP providers find it easy to support PrEP users to adhere to their chosen regimen because nationally-developed patient information booklets with key points about the various ways to take PrEP and diagrams showing how to follow event-based PrEP aid provision of accurate dosing advice | **Example 1**  “*I don’t know how good I would be if they were saying so I’m going to have sex on a Saturday and then I’m going to have sex on a Thursday, when do I actually start and stop it, you know. So, it’s case-by-case and I probably still need to refresh my memory a little bit and read up a bit on that still if I was doing that because most of the people are just taking it every day*.” (Sexual healthcare professional)  **Example 2**  “*Having something like a patient information leaflet just allows you to codify your advice very clearly which is actually more useful to clinicians than we give credit for*.” (Sexual healthcare professional) | Knowledge  Memory, attention, and decision processes  Environmental context and resources | Education  Enablement | 4.1 Instruction on how to perform a behaviour  7.1 Prompts/cues  2.7 Feedback on behaviour | 22. Use a multi-method approach to educate sexual healthcare professionals about event-based dosing (4.1) and assist them during consultations (7.1). For example, include clear written instructions and diagrams that depict how to take event-based PrEP, including examples of when to start and stop for various scenarios, in a range of resources (e.g. brief fact sheet, PrEP provider pocket guide, national patient information booklets), provide sexual healthcare professionals with laminated copies of the event-based dosing diagrams that they can pin to their wall as a quick reminder of how to use event-based PrEP, record a short video or soundbite that explains event-based dosing for different scenarios that sexual healthcare professionals may watch or listen to at a future date, include an online or paper-based quiz with questions about event-based dosing as part of sexual healthcare professionals PrEP training and ongoing CPD and ensure that there is sufficient time or a named person to contact to discuss the answers after, if necessary (2.7) | 22. Keep but modify (discussed) – lots of nice examples to provide detail to headline recommendations. Consider who hosts the resources/ joined up approach/ ensure not developed piece-meal in different regions at same time. Need user involvement to tailor resources to key populations and ensure issues of relevance are covered. | PrEP services should use a joined-up, multi-method approach to improve PrEP providers’ understanding of event-based dosing to assist them during consultations. The following approaches could help: a range of resources (e.g. national, co-produced PrEP provider pocket guide and patient information, short videos, wall-mounted displays) with clear written instructions and diagrams depicting correct usage of event-based PrEP, including examples of when to start and stop for various scenarios, and a quiz with questions about event-based dosing as part of PrEP training |

**Table S2: Priority area 2- A BCW analysis of ‘PrEP users consistently take PrEP appropriately’**

| **Barriers** | **Facilitators** | **Indicative quotes** | **TDF domains** | **Intervention Functions** | **Potential BCTs**  **Numbers = BCTs** | **Recommendations for those considering implementing PrEP at scale**  **Accept/Reject/Modify**  **Numbers in brackets = BCTs** | **Post APEASE and expert input decision**    **Accept/Reject/Modify** | **Agreed final recommendations for those considering implementing PrEP at scale** |
| --- | --- | --- | --- | --- | --- | --- | --- | --- |
| PrEP users find it difficult to consistently take PrEP appropriately because of the absence of, or disruption to, a daily or usual routine (including having spontaneous sex while using event-based PrEP) | PrEP users find it easy to consistently take PrEP appropriately because they incorporate  taking PrEP into a pre-existing daily routine (e.g. having breakfast, brushing teeth, taking other medication or vitamins if taking PrEP once a day) or a usual routine ahead of planned sex (if using event-based PrEP) | “*I do take medication already, daily, other mediation, so it's just an extra tablet in the morning. If it had been the more complicated dosage, of like, you know, two tables, and doing one and one, I would probably get more confused. But the one daily fits in pretty well with my lifestyle*.” (PrEP user) | Memory, attention and decision processes  Environmental context and resources  Behavioural regulation | Environmental restructuring  Enablement | 7.1 Prompts/cues  1.4 Action planning  2.3 Self-monitoring of behaviour | 41. Advise PrEP users to formulate an ‘if-then’ plan that links taking PrEP once a day to a specific task which remains constant even in the absence of or disruption to a daily routine (1.4, 7.1)  41. Suggest that PrEP users mark off on a calendar or record in a diary whether they have taken their daily medication (2.3)  Sexual healthcare professionals, other HCPs providing PrEP care, and NGO staff should share this practical tip with PrEP users. Practical tips should also appear in national patient information booklets and online resources (e.g. sexual health services, NGO, and HIV/PrEP activists’ websites and social media) | 41. Keep (discussed) – merge with others marked 41.  Rec merged with others marked 41 including those in priority area 1 ‘PrEP providers support PrEP users to adhere to their chosen regimen’. Did not include that final rec in this priority area after de-duplication (felt more appropriate in priority area 1 and to include the final rec to the right in this priority area instead)  41. Keep (discussed) – merge with others marked  41.  Rec merged with others marked 41 including those in priority area 1 ‘PrEP providers support PrEP users to adhere to their chosen regimen’. Did not include that final rec in this priority area after de-duplication (felt more appropriate in priority area 1 and to include the final rec to the right in this priority area instead) | PrEP users should consider a range of strategies, including those outlined in priority area one, to ensure effective use of PrEP and share those they find beneficial with potential and other PrEP users |
| PrEP users find it difficult to consistently take PrEP appropriately because rigid clinic appointment processes mean it is challenging to access PrEP ‘last minute’ so users can run low on or out of PrEP | -- | “*The difficulty is where you have DNAs or people just choosing to come to the walk-in clinic for follow-up PrEP and the nursing team not being in a position to be able to do that and being able to manage patient expectations in terms of that consultation. And so it’s about trying to reinforce with the patient that the follow-up is at these dedicated times, certainly with their agreement, but they have to come to those appointments for their further prescriptions to be given, you can’t just pop up on the off-chance that you’ll be given a further supply of PrEP. So it’s been about that, managing the DNAs and then trying to fit them in somewhere else and already stretched clinics and them saying they’re running out of medication and then you feeling duty bound to try your best, to try and ensure they don’t have gaps in the provision of the medication*.” (Sexual healthcare professional) | Memory, attention and decision processes  Environmental context and resources | Environmental restructuring  Enablement | 12.1 Restructure the physical environment | 00. Sexual healthcare professionals should support PrEP users (3.2) to navigate services for appropriate expert support. Support could include providing clear information on how to get further PrEP prescriptions (i.e. clinic-specific processes, managing expectations - PrEP not an emergency, try and plan appointments in advance as clinics can fill up quickly)  2. Establish PrEP as routine clinical practice within sexual health services and implement PrEP monitoring through regular drop-in clinics, in addition to booked appointments (12.1) | 2. Keep (discussed) – flexible service for those whom fixed appointments do not suit. Will improve access to reviews but could be issues re: staff competencies and time, especially if there are things on “shopping list”, such as symptoms of STIs. *  Duplicate and merged with 4 and 6a which also relate to flexible provision of individualised PrEP care that meets diverse needs – included as a final rec for PA3 ‘PrEP users attend PrEP reviews’ (as 4 and 6a relate to this PA too) rather than for this PA. | PrEP providers and NGO staff should support PrEP users to navigate services and online information for appropriate expert support. *Support could include: providing clear information on how to get further PrEP prescriptions (i.e. clinic-specific processes, managing expectations - PrEP not an emergency, try and plan appointments in advance as clinics can fill up quickly); ensuring PrEP users know they can return to or call the PrEP service for adherence support and have the option to change regimens; and raising awareness of and directing PrEP users to reputable online sources of adherence support* |
| **--** | PrEP users find it easy to consistently take PrEP appropriately because they receive adherence support from sexual healthcare professionals (e.g. at PrEP appointments, through provision of nationally-developed patient information booklets, on an ad-hoc basis) | **Example 1**  “*The first question, it's looking at adherence, have they had any side-effects, or have they managed to take it, are they remembering it every day. Or if its event based, are they remembering to take it as they should. Is event based still the thing for them, do they want to change onto daily*.” (Sexual healthcare professional)  **Example 2**  “*It was quite a visual leaflet…it would give almost like a timeline of how it would work, and it showed very clearly what the difference was between event based and also daily dosing, that was really, really helpful*. (PrEP user)  **Example 3**  “*…keep the leaflet there so you can refer to it and what exactly you need to do and if you’ve got any problems, give us a phone. We always give out the details of the clinic for phoning for any queries*.” (Sexual healthcare professional) | Environmental context and resources  Professional role and identity  Social influences | Environmental restructuring  Education  Persuasion  Enablement | 7.1 Prompts/cues  5.1 Information about health consequences  9.1 Credible source  4.1 Instruction on how to perform the behaviour  1.1 Goal setting (behaviour)  1.4 Action planning  1.2 Problem solving  3.1 Social support (unspecified)  1.5 Review behavioural goal(s) | 16. Create paper-based or electronic checklists/proformas (based on formal protocols for PrEP initiation and monitoring) that prompt sexual healthcare professionals to cover adherence-related issues at all PrEP appointments (7.1)  42. Sexual healthcare professionals must educate PrEP users on the importance of good adherence to ensure PrEP efficacy and minimise risks of HIV infection and antiretroviral resistance (5.1, 9.1)  43. Sexual healthcare professionals should provide PrEP users with verbal, written, and visual instructions re: medication dose, schedule, lead-in time to protection, and missed doses for the various ways of taking PrEP (e.g. via national patient information leaflet, wallet-sized insert) (4.1). Ensure these materials are also available in clinic waiting areas and at other relevant settings (e.g. NGOs)  44. At PrEP initiation, sexual healthcare professionals could set PrEP users a goal in terms of the behaviour to be achieved (e.g. daily dosing) (1.1, 1.4) and engage them in coping planning (1.2) to overcome barriers / increase facilitators to taking PrEP as per their chosen regimen. During PrEP monitoring, sexual healthcare professionals should explore how well PrEP users have adhered to their chosen regimen (1.5) and with their agreement, either reset the same goal (i.e. stick to same regimen) or modify the future goal (i.e. switch regimen) (1.1)  45. Sexual healthcare professionals could direct PrEP users to reputable online sources of adherence support (e.g. sexual health services, NGO and HIV/ PrEP activists’ websites and social media) (3.1, 9.1) in addition to the information they provide (e.g. verbally, via provision of national patient information booklet)  37. Sexual healthcare professionals should inform PrEP users how to access the sexual health service for ad-hoc adherence support between appointments and ensure contact details and opening hours are kept up to date on the sexual health service website (3.1) | 16. Keep (discussed) – could be an example of how to support adherence (relates to safe-care and people feeling that protocols are a good way of achieving this). Could be combined with other recommendations re: protocols.  42. Keep (discussed) – to be merged with other adherence-related recommendations.  43. Keep (discussed) – to be merged with other adherence-related recommendations.  44. Keep but modify (discussed) – use professional judgement to decide whether an explicit exercise in goal setting and coping planning is required.  45. Keep (discussed) – flexible services that meet local population needs? Merged with 00 and 37 to create the final rec.  37. Keep but modify (discussed) – don’t want to encourage people to have ad-hoc appointments but people do need the confidence to navigate the healthcare system (i.e. know they can come back to speak to a sexual healthcare professional, option to change regimens). Merged with 00 and 37 to create the final rec. Duplicate. | PrEP services should create checklists/proformas, based on formal protocols, to prompt PrEP providers to cover adherence-related issues during PrEP initiation and reviews  PrEP providers should emphasise the importance of adherence to minimise risks of HIV transmission or antiretroviral resistance and provide verbal, written, and visual instructions regarding medication dose, schedule, lead-in time to protection, and missed doses for the different ways of taking PrEP  PrEP providers should consider offering PrEP users an explicit exercise in goal setting, coping planning, and review of behavioural goals to support adherence to their chosen PrEP regimen  PrEP providers and NGO staff should support PrEP users to navigate services and online information for appropriate expert support. *Support could include: providing clear information on how to get further PrEP prescriptions (i.e. clinic-specific processes, managing expectations - PrEP not an emergency, try and plan appointments in advance as clinics can fill up quickly); ensuring PrEP users know they can return to or call the PrEP service for adherence support and have the option to change regimens; and raising awareness of and directing PrEP users to reputable online sources of adherence support* |
| **--** | PrEP users find it easy to consistently take PrEP appropriately because they put in place reminders to avoid missing a dose (e.g. phone alarm or alert on an app, use of a pill organiser) | “*When your phone buzzes at 12 o'clock then you know it's time to take your pill. I've found that helps*.” (PrEP user)  *“I decided to use a dosette box and have all of my medications there and that’s kind of keeping me in check of taking them daily.”* (PrEP user) | Memory, attention and decision processes  Environmental context and resources  Behavioural regulation | Environmental restructuring  Enablement | 7.1 Prompts/cues  12.5 Adding objects to the environment | 41. Encourage PrEP users (e.g. during interactions with sexual healthcare professionals and NGO staff, via sexual health services, NGO, and HIV/PrEP activists’ websites and social media, in national patient information booklets) to set up reminder alarms (7.1) and/or use a pill organiser (7.1, 12.5) as prompts to take PrEP | 41. Keep (discussed) – merge with others marked 41.  Rec merged with others marked 41 including those in priority area 1 ‘PrEP providers support PrEP users to adhere to their chosen regimen’. Did not include that final rec in this priority area after de-duplication (felt more appropriate in priority area 1 and to include the final rec to the right in this priority area instead) | PrEP users should consider a range of strategies, including those outlined in priority area one, to ensure effective use of PrEP and share those they find beneficial with potential and other PrEP users |
| **--** | PrEP users find it easy to consistently take PrEP appropriately because they keep PrEP handy by carrying it on them (e.g. in a bag, jacket pocket) and storing it in convenient places around and outside the home (e.g. in the car, at work) | **Example 1**  *“I keep it somewhere where I look through, like in my backpack, where I would look for many things…then if I forget, I will very soon see it, as in, see the box with the PrEP and be like, oh did I take it, oh yeah I did, or, oh no I didn’t.”* (PrEP user)  **Example 2**  “*I would actually have one bottle in my bag, I’d have one in the living room and one in the bedroom so wherever I was during the day I could actually take it..because sometimes I’d remember and I’d think, oh, wait a minute, and if I was at work I could take it if I needed to. Because I’d have one in my bag*.” (PrEP user) | Memory, attention and decision processes  Environmental context and resources  Behavioural regulation | Environmental restructuring  Enablement | 12.1 Restructure the physical environment  7.1 Prompts/cues | 41. Advise PrEP users to keep PrEP handy by carrying it on them and storing it in convenient places around and outside the home (12.1), both as a prompt to take PrEP (7.1) and to ensure it is readily accessible  Sexual healthcare professionals, other HCPs providing PrEP care, and NGO staff should share this practical tip with PrEP users. Practical tips should also appear in national patient information booklets and online resources (e.g. sexual health services, NGO, and HIV/PrEP activists’ websites and social media) | 41. Keep (discussed) – merge with others marked 41.  Rec merged with others marked 41 including those in priority area 1 ‘PrEP providers support PrEP users to adhere to their chosen regimen’. Did not include that final rec in this priority area after de-duplication (felt more appropriate in priority area 1 and to include the final rec to the right in this priority area instead) | PrEP users should consider a range of strategies, including those outlined in priority area one, to ensure effective use of PrEP and share those they find beneficial with potential and other PrEP users |

### Table S3 - Priority area 3- A BCW analysis of ‘PrEP users attend PrEP reviews’

| **Barriers** | **Facilitators** | **Indicative quotes** | **TDF domains** | **Intervention Functions** | **Potential BCTs**  **Numbers = BCTs** | **Recommendations for those considering implementing PrEP at scale**  **Accept/Reject/Modify**  **Numbers in brackets = BCTs** | **Post APEASE and expert input decision**    **Accept/Reject/Modify** | **Agreed final recommendations for those considering implementing PrEP at scale** |
| --- | --- | --- | --- | --- | --- | --- | --- | --- |
| PrEP users find it difficult to attend PrEP reviews because there are limited options for where (e.g. at some not all sexual health services, located far away), when (e.g. inconvenient time slots), and how (e.g. by appointment, set up to be delivered in male only or GBMSM clinics) they can access them | PrEP users find it easy to attend PrEP reviews because there is flexibility in where (e.g. at all sexual health services, in other more valued / acceptable settings), when (e.g. extended opening hours), and how (e.g. via drop-in clinics, by appointment) they can access them | **Example 1**  *“They can't take the kidney tests in the [outreach] clinic that’s dedicated to gay men, because it's in a different venue…so, essentially, if at those clinics, if they could take the kidney test as well.”* (PrEP user)  **Example 2**  “*Because we’re a rural area, we might have patients that live a distance away, and with work commitments, it might make it difficult for them to come into the clinic. Whereas if they live a distance away, but they happen to be in [town], then on a particular day, if they were able to just get it from a drop-in clinic, then it would be easier for them.*” (Sexual healthcare professional) | Environmental context and resources | Environmental restructuring  Enablement | 12.1 Restructure the physical environment  12.2 Restructure the social environment  3.1 Social support (unspecified) | 1. Consider alternative service models to make PrEP monitoring available to PrEP users via a range of settings, including all sexual health services (e.g. local hubs and satellites, as well as central services), remote care (e.g. ePrEP, phone consultations), community venues (e.g. outreach clinics), and non-sexual health-specific health services (e.g. reproductive health clinics, GP surgeries), with agreed pathways for non-complex PrEP users and those with additional medical complexity (12.1, 12.2)  2. Establish PrEP as routine clinical practice within sexual health services and implement PrEP monitoring through regular drop-in clinics, in addition to booked appointments (12.1)  3. Maximise all drop-in visits by ensuring there is sufficient waiting space, toilets, and consultation rooms (12.1) and operationalising drop-in clinics via a multidisciplinary team of sexual healthcare professionals who can task-share and accommodate complex cases (12.2)  4. Provide access to drop-in clinics and pre-bookable appointments on mid-week evenings and at weekends to suit contemporary lifestyles and meet local population needs (12.1)  5. Support PrEP users in becoming aware of when and how they can access drop-in clinics and book and reschedule appointments for PrEP monitoring (e.g. sexual healthcare professionals provide information verbally, hand out location-specific leaflets or wallet-sized inserts, signpost to websites) (3.1) | 1. Keep but modify (discussed) – there has to be reasonable provision of PrEP throughout the wider healthcare system to extend reach, as otherwise some people will not benefit.  Increased reach of PrEP could be viewed as aspirational. Too much detail at the moment, lose from ‘including all sexual health services’ to ‘GP surgeries’. Some concerns are that HCPs in non-specialist settings may not see many PrEP users, effectiveness, cost and monitoring, equity in terms of the digital divide. See also, notes for 62 in stage 1 (said too vague). *  2. Keep (discussed) – flexible service for those whom fixed appointments do not suit. Will improve access to reviews but could be issues re: staff competencies and time, especially if there are things on “shopping list”, such as symptoms of STIs. * Duplicate. Merged with 4 and 6a which also relate to flexible provision of individualised PrEP care that meets diverse needs.  3. Reject (discussed) – what services should be doing anyway and an underlying assumption about what is available in order to support delivery of good SRH in general. Not specific to PrEP.  4. Keep (discussed) – relates to flexible service provision. * Merged with 2 and 6a (see 2 for the final rec).  5. Reject (discussed) – too general and already part of existing service. | PrEP commissioners should consider offering established PrEP users reviews in a range of settings (not solely sexual health clinics). *Each service model should incorporate pathways for non-complex PrEP users and those with additional medical complexity*  PrEP services should ensure flexible provision of individualised PrEP care that meets diverse needs. *Examples include: implementing PrEP reviews through drop-in clinics as well as booked appointments (as the programme matures); providing evening and weekend access to suit lifestyles and meet local population needs; ensuring there are options for how to book in for the next review (e.g. online, by phone, in-person), with the appointment system open far enough in advance to enable booking in before leaving the premises; and flexibility to provide extra PrEP supply to accommodate longer periods between reviews, if necessary*  -- |
| PrEP users find it difficult to attend PrEP reviews because of an absence of appointment scheduling, reminder, follow-up, and/or other targeted intervention processes | PrEP users find it easy to attend PrEP reviews because there are appointment scheduling, reminder, follow-up, and/or other targeted intervention processes in place | **Example 1**  *“Once you’re finished and got the PrEP you actually just make the appointment for the next three months at that point, and then you go back and you go through the tests again…”* (PrEP user)  **Example 2**  “*They send a text the following day after the appointment’s made, and then they send a text two days prior to the appointment to confirm your time. So that works really well*.” (PrEP user)  **Example 3**  “*If they have DNA’d the appointment we may give them a follow-up phone call, but if they have just not made another appointment then we don't follow them up*.” (Sexual healthcare professional) | Environmental context and resources  Memory, attention and decision processes  Behavioural regulation | Environmental restructuring  Enablement | 12.1 Restructure the physical environment  7.1 Prompts/cues  2.2 Feedback on behaviour  3.1 Social support (unspecified) | 6a. Ensure the appointment system is open far enough in advance to enable PrEP users to book their next PrEP appointment before leaving the premises (12.1)  7. Prompt sexual healthcare professionals (e.g. via paper-based or electronic checklists/ proformas, SOPs, ‘pop-up’ messages within the IT system) to remind PrEP users to book their next appointment before leaving the premises (7.1)  8. Use an automated reminder system to alert PrEP users (e.g. via email, voice message, SMS) to a booked appointment (7.1) or to notify that they are overdue to attend (2.2)  9. Integrate ‘pop-up’ messages into the IT system to inform sexual healthcare professionals that a PrEP user did not attend or are overdue a PrEP appointment and advise exploration of the issue (7.1)  10a. Run a monthly report on the IT system to identify ‘did not attends’ and those overdue a PrEP appointment and attempt to make contact with PrEP users (e.g. via email, SMS, phone) and reengage them with PrEP care, if appropriate (3.1) | 6a. Keep but modify (discussed) - Doesn’t need to be done physically at the time, could be done remotely. Key point is to ensure availability of next slot at required time and ability to “flex” the timing if necessary. For example, extra PrEP supply if away somewhere or unsure. May not suit those who have less fixed plans. Does make it easier to follow up defaulters. Merged with 2 and 4.  7. Reject (discussed) – Reject all pop-ups. Already happens. Duplicate.  8. Keep but modify (not discussed) – may not be acceptable for everyone so need to have opt-out. Need reminder in advance of an upcoming review and further prompt if overdue.  9. Reject (discussed) – favour automated system which sends reminders to PrEP users. Duplicate.  10a. Keep (discussed) – about knowing your PrEP cohort (at service-level). Findings can inform PrEP initiation process (develop interventions for those who miss appointments/ are more likely to not reattend  improve retention rates). | See 2 for the final rec  --  PrEP services should use existing or introduce new clinic processes, such as an automated text message system (with opt-out option, to remind and follow-up PrEP users about PrEP reviews and to try and reengage non-attenders.  --  PrEP services should research their patient cohort and available literature to identify characteristics of people likely to miss appointments or not re-attend for PrEP reviews and develop interventions to be delivered at PrEP initiation to improve retention |
| PrEP users find it difficult to attend PrEP reviews because they do not require a PrEP prescription (e.g. they are doing event-based dosing or have stopped PrEP in the interim period) | PrEP users find it easy to attend PrEP reviews because they value the regular sexual health screening and other health tests and discussions that take place within PrEP reviews | **Example 1**  “*It's sometimes a struggle to get them back at three months for review. What brings them back essentially is wanting more medication and actually if they don't want more medication they're less inclined to return for review, and obviously if they've had any risky sexual behaviour then they should be getting an STI screen. So it can be difficult to get those men back*.” (Sexual healthcare professional)  **Example 2**  “*If you’re constantly getting kidney and liver function tests and it comes back positive, then everything’s working fine…so, that kind of reassures me about my health. I think they’re very important*.” (PrEP user) | Beliefs about consequences  Behavioural regulation | Education  Persuasion  Enablement | 5.1 Information about health consequences  5.6 Information about emotional consequences  9.1 Credible source  1.8 Behavioural contract  1.9 Commitment  1.1 Goal setting (behaviour) | 31a. Provide PrEP users with a range of information sources (e.g. posters, national patient information booklets, positive testimonials of PrEP users, online resources, verbal communication by sexual healthcare professionals and NGO staff) regarding the health and emotional benefits of PrEP monitoring, including the importance of regular HIV and STI testing and discussing stopping PrEP with a sexual healthcare professional (5.1, 5.6, 9.1)  01. Sexual healthcare professionals could ask PrEP users to verbally agree to or sign a written contract specifying that they will attend for regular PrEP reviews, even if they are doing event-based dosing or have stopped PrEP in the interim period (1.8, 1.9, 1.1) | 31a. Keep but modify (discussed) –easy/inexpensive though not PrEP specific, would apply to any chronic condition, and will have a small diminishing effect. Ideally info should be co-produced and tailored for different populations. Also include the importance of renal monitoring and review of how things are going. One resource for use across PrEP continuum.  01. Reject-HP only input -impractical and too paternalistic. regular reviews is softer and relates to the idea of good PrEP citizenship. | PrEP providers and NGO staff should encourage good PrEP citizenship by emphasising the health and emotional benefits of PrEP reviews, such as regular HIV and STI testing, renal monitoring and review of ‘how things are going’, and the importance of discussing stopping PrEP with a PrEP provider. Information sources may include co-produced patient information and verbal communication  PrEP users should commit to attending regular PrEP reviews, even if they do not require a new PrEP prescription when the next review is due |
| -- | PrEP users find it easy to attend PrEP reviews because PrEP providers are explicit about the requirement for PrEP monitoring at the outset | “*They kind of make this agreement. So, if you want to be on PrEP funded by the NHS, this is the expectation …you come every three months and we do this, this and this and if you don’t come every three months, if you miss your appointments, you may fall off the protocol to still be funded for PrEP. So, it’s very much like an agreement. So, all that is definitely explained and set out to them at the initial assessment*.” (Sexual healthcare professional) | Reinforcement  Behavioural regulation | Coercion  Enablement | 10.11 Future punishment  1.8 Behavioural contract  1.9 Commitment  1.1 Goal setting (behaviour) | 32. Sexual healthcare professionals should inform PrEP users at the initial assessment that they risk losing their access to PrEP if they do not attend for regular PrEP reviews (10.11)  01. Sexual healthcare professionals could ask PrEP users to verbally agree to or sign a written contract specifying that they will attend for regular PrEP reviews, even if they are doing event-based dosing or have stopped PrEP in the interim period (1.8, 1.9, 1.1) | 32. Reject (not discussed) – too paternalistic and not person-centred. Feels punitive and controlling. Strong negative reaction to this one.  01. Reject-HP only input -impractical and too paternalistic. regular reviews is softer and relates to the idea of good PrEP citizenship. | Do not take forwards  Do not take forwards |

### Table S4 - Priority area 4- A BCW analysis of ‘PrEP providers reassess PrEP users’ candidacy based on risk of HIV acquisition’

| **Barriers** | **Facilitators** | **Indicative quotes** | **TDF domains** | **Intervention Functions** | **Potential BCTs**  **Numbers = BCTs** | **Recommendations for those considering implementing PrEP at scale**  **Accept/Reject/Modify**  **Numbers in brackets = BCTs** | **Post APEASE and expert input decision**    **Accept/Reject/Modify** | **Agreed final recommendations for those considering implementing PrEP at scale** |
| --- | --- | --- | --- | --- | --- | --- | --- | --- |
| PrEP providers find it difficult to reassess PrEP users’ candidacy based on risk of HIV acquisition because they overlook this aspect of PrEP monitoring (e.g. familiarity and routinisation of giving out PrEP, assume PrEP users have an ongoing need) | PrEP providers find it easy to reassess PrEP users’ candidacy based on risk of HIV acquisition because supporting documents and the IT system prompt them to undertake this task | **Example 1**  “*The danger to that is, because* *you can get a bit complacent about it and think that this is just doing tests and handing out drugs, and not properly reviewing people… checking that they still fit the eligibility criteria, and things like that*.” (Sexual healthcare professional)  **Example 2**  *“They didn’t really go into depth to see if I was still eligible. They kind of assumed.”* (PrEP user)  **Example 3**  “*She created template documents for us to use that would prompt us to ask the right questions, consider the right things, and we had one for initial assessment, we had one for a one-month review and we had one for a three-month review*.” (Sexual healthcare professional) | Memory, attention and decision processes  Environmental context and resources  Behavioural regulation | Environmental restructuring  Enablement  Persuasion | 7.1 Prompts/cues  2.3 Self-monitoring of behaviour  2.4 Self-monitoring of outcome(s) of behaviour  5.1 Information about health consequences  9.1 Credible source | 11. Create paper-based or electronic checklists/ proformas (based on a formal protocol for PrEP monitoring) that prompt sexual healthcare professionals to reassess PrEP users’ eligibility (7.1) and request documentation of their eligibility decision (2.3) with a record of the reasons to support it (2.4)  12. Introduce interactive ‘pop-up’ messages within the IT system (7.1) that alert sexual healthcare professionals to the potential health risks of PrEP (5.1) and require them to confirm they have reassessed PrEP users’ eligibility (2.3) prior to prescribing  17. Use a multi-method approach to educate sexual healthcare professionals about the importance of reassessing PrEP users’ eligibility to avoid them taking PrEP unnecessarily if they are no longer at high risk for HIV acquisition (5.1, 9.1) | 11. Reject (discussed) – could help those who may wish to consider stopping PrEP or changing regimen. But tick boxes often dehumanise and impair proper engagement and discussion plus more concern that PrEP would be removed from people still at risk since current eligibility criteria /risk assessments aren’t sensitive enough for all key populations.  12. Reject (not discussed) – staff hate pop-ups.  17. Reject (discussed) – basic competence. Also, as PrEP users move between initiation (face to face) to follow-up (remote?), its perhaps better to have annual check ins where these issues can be addressed. | Do not take forwards  Do not take forwards  Do not take forwards |

**Table S5 Priority area 5 A BCW analysis of ‘ PrEP providers address wider sexual health issues’**

| **Barriers** | **Facilitators** | **Indicative quotes** | **TDF domains** | **Intervention Functions** | **Potential BCTs**  **Numbers= BCTs** | **Recommendations for those considering implementing PrEP at scale**  **Accept/Reject/Modify**  **Numbers in brackets=BCTs** | **Post APEASE and expert input decision**    **Accept/Reject/Modify** | **Agreed final recommendations for those considering implementing PrEP at scale** |
| --- | --- | --- | --- | --- | --- | --- | --- | --- |
| PrEP providers find it difficult to address wider sexual health issues because of the time constraints of PrEP review appointments | PrEP providers find it easy to address wider sexual health issues because they have generous and/or flexible appointment times for PrEP reviews | **Example 1**  *“These can potentially be quite lengthy and complex dialogues that aren't necessarily going to be able to be accommodated within a short consultation on a three-monthly basis.”* (NGO staff)  **Example 2**  “*In [urban Health Board], I know they're really pushed for time in the PrEP clinics, whereas here, we are a bit more flexible, and we can kind of, we have time to chat as much or as little as they want*.” (Sexual healthcare professional) | Environmental context and resources  Professional role and identity | Environmental restructuring  Education  Persuasion  Enablement | 12.2 Restructure the social environment  5.1 Information about health consequences  5.3 Information about social and environmental consequences  5.6 Information about emotional consequences  2.3 Self-monitoring of behaviour  2.4 Self-monitoring of outcome(s) of behaviour  2.2 Feedback on behaviour  2.7 Feedback on outcome(s) of behaviour  3.1 Social support (unspecified) | 13. Sexual health services should explore and provide innovative ways of scheduling appointments with built-in flexibility to respond to long standing health inequalities in health and HIV literacy and varying needs of PrEP users (e.g. longer discussions about PrEP and wider sexual health issues) (12.2)  23. Facilitate and sustain an organisational culture that values a wholistic approach to sexual health and wellbeing (12.2) (e.g. reflect a wholistic approach in the sexual health service values and mission statement and include as a core competency for professional conduct, address in education sessions (5.1, 5.3, 5.6) and reflective practice, and as part of annual appraisals (2.3, 2.4, 2.2, 2.7))  27. Check that PrEP users are aware of other specialist services available locally (e.g. delivered by NGOs, available within the sexual health service) and signpost or make a direct referral, as necessary (3.1) | 13. Keep but modify (discussed) – not PrEP specific but providing individualised PrEP care and responding to long standing health inequalities in health and HIV literacy and varying need is important and needs to feature somewhere (flexibility of service provision). ‘Innovative’ is too subjective. Duplicate.  23. Reject (not discussed) – don’t dispute its importance, but favour other PrEP-specific recommendations. This should already be embedded in good clinical practice. Duplicate  27. Keep but modify (discussed) – reciprocity of connection that is in stage 1. Duplicate. | PrEP services should ensure flexible provision of individualised PrEP care that meets diverse needs. For example, explore and provide ways of scheduling appointments with built-in flexibility to respond to long-standing inequalities in health and HIV/PrEP literacy during consultations  PrEP providers and NGO staff should support PrEP users to navigate services and online information for appropriate expert support. *Support could include signposting and/or referring PrEP users to other specialist services across and beyond the HIV prevention and care sector, as necessary* |
| -- | PrEP providers find it easy to address wider sexual health issues because they have built trusting relationships and familiarity with PrEP users through continuity of care | “*I certainly feel the…you know, the advantage to it, because you know them and they feel comfortable to tell you things and you feel comfortable to ask them things and you, kind of, know what’s been going on. You know, because, you saw them last time, so you know the questions that you asked ...and you, kind of, pick up where you left off*.” (Sexual healthcare professional) | Environmental context and resources  Social influences  Professional role and identity | Environmental restructuring  Education  Modelling  Enablement | 12.2 Restructure the social environment  12.1 Restructure the physical environment  7.1 Prompts/cues  5.1 Information about health consequences  5.3 Information about social and environmental consequences  6.1 Demonstration of the behaviour  2.2 Feedback on behaviour  2.3 Self-monitoring of behaviour | 14. Where possible, assign each PrEP user a ‘usual' sexual healthcare professional and operate a buddy system where paired sexual healthcare professionals can see each other's patients, for example, when the other is on leave, to facilitate continuity of care (12.2)  6b. Ensure the appointment system is open and the rota agreed far enough in advance to enable PrEP users to book their next PrEP appointment with their ‘usual’ sexual healthcare professional or buddy before leaving the premises (12.1)  7. Prompt sexual healthcare professionals (e.g. via paper-based or electronic checklists/ proformas, SOPs, ‘pop-up’ messages within the IT system) to remind PrEP users to book their next appointment before leaving the premises (7.1)  24. Facilitate and actively maintain (e.g. via training, huddles, clinical supervision, reflective practice) a warm, welcoming, and friendly atmosphere wherein sexual healthcare professionals communicate with patients in a non-judgemental manner, using inclusive, sex- and PrEP-positive, and destigmatising language to establish trust and ensure an open dialogue (12.2, 5.3)  18. Promote the advantages of high-quality clinical record keeping for continuity of care (5.1, 5.3), share best practice examples that meet the standards set out by the sexual health service and/or relevant professional bodies (6.1), and appraise and encourage sexual healthcare professionals to reflect on their sRejects of recording episodes of care (2.2, 2.3) | 14. Reject (not discussed) – totally impractical in the real world. Also would limit training opportunities and foster dependence. Duplicate.  6b. Reject (not discussed) – impractical for PrEP users to have a designated sexual healthcare professional for reviews. The bit about the appointment system being open in advance is covered in 6a. Duplicate.  7. Reject (discussed) – Reject all pop-ups. Already happens. Duplicate.  24. Reject (discussed) – support the general sentiment but is not PrEP specific. Useful content for the intro as we will need to make a statement pointing towards existing standards / expectations of the bedrock of delivery. Duplicate.  18. Reject (not discussed) – not PrEP specific. Is addressed in existing clinical governance, appraisal and revalidation. Duplicate | --  --  --  --  -- |
| -- | PrEP providers find it easy to address wider sexual health issues because they are trained to deliver brief behaviour change interventions or have the option to signpost PrEP users and/or make direct referrals to other specialist services (e.g. for drug and alcohol problems, gender clinic, rape/sexual assault) for appropriate support | “*It definitely feels better than it did years and years ago when I started. It was a bit like I don’t even know what to say now. I don’t even know. You know, you told me this awful thing and I feel I want to have an answer for you and I don’t know what the answer is. So, I do feel like there’s more accessible support that you could refer someone to*.” (Sexual healthcare professional) | SRejects  Professional role and identity  Environmental context and resources | Training  Modelling  Environmental restructuring  Enablement | 4.1 Instruction on how to perform the behaviour  6.1 Demonstration of the behaviour  8.1 Behavioural practice/rehearsal  2.2 Feedback on behaviour  2.3 Self-monitoring of behaviour  12.2 Restructure the social environment  3.1 Social support (unspecified) | 19. Develop sexual healthcare professionals’ sRejects in delivering brief behaviour change interventions through interactive activities (e.g. via workshops, online courses, in clinical supervision), including training on the technical aspects (4.1), video examples and shadowing of more experienced sexual healthcare professionals for ‘what works’ tips (6.1), role-/real-play exercises (8.1) with provision of feedback (2.2), and ongoing reflections on sReject acquisition (2.3)  25. Establish good connections with other specialist services (e.g. delivered by NGOs, those available within the sexual health service) (12.2) that sexual healthcare professionals could signpost and/or directly refer PrEP users to, for appropriate expert support (3.1)  26. Develop and raise sexual healthcare professionals’ awareness of protocols to ensure standardisation in care navigation (e.g. signposting and referrals to other specialist services) (4.1) | 19. Reject (not discussed) – not PrEP specific. Support in general but for broader delivery of quality care.  25. Keep but modify (discussed) – Two-way connections / partnership work but need to word carefully so doesn’t seem like sexual health services are passing the buck and being mindful of 3^rd^ sector funding cuts. Include NGOs that serve communities other than GBMSM. Duplicate  26. Reject (not discussed) | --  PrEP services and NGOs should enhance and maintain good connections across HIV prevention and care and other specialist services, to facilitate easy reciprocal referrals. Consider carefully the type of support required and which service is best placed to provide it  -  Not taken forwards |

### Table S6 Priority area 6- A BCW analysis of ‘PrEP users discuss wider sexual health issues’

| **Barriers** | **Facilitators** | **Indicative quotes** | **TDF domains** | **Intervention Functions** | **Potential BCTs**  **Numbers= BCTs** | **Recommendations for those considering implementing PrEP at scale**  **Accept/Reject/Modify**  **Numbers in brackets=BCTs** | **Post APEASE and expert input decision**    **Accept/Reject/Modify** | **Agreed final recommendations for those considering implementing PrEP at scale** |
| --- | --- | --- | --- | --- | --- | --- | --- | --- |
| PrEP users find it difficult to discuss wider sexual health issues because PrEP reviews feel rushed and are typically focused on PrEP only | -- | “*They don’t really say, well, you know, what’s your…what are you currently up to? Are you seeing anyone or…you know, there’s no, kind of, counselling service, if that makes… if that’s the right term to use. There’s no, kind of, how are you in your life and how are you within your sexual health, kind of thing. There’s none of that at all*.” (PrEP user) | Environmental context and resources  Professional role and identity | Environmental restructuring  Education  Persuasion  Enablement | 12.2 Restructure the social environment  5.1 Information about health consequences  5.3 Information about social and environmental consequences  5.6 Information about emotional consequences  2.3 Self-monitoring of behaviour  2.4 Self-monitoring of outcome(s) of behaviour  2.2 Feedback on behaviour  2.7 Feedback on outcome(s) of behaviour  3.1 Social support (unspecified) | 13. Sexual health services should explore and provide innovative ways of scheduling appointments with built-in flexibility to respond to long standing health inequalities in health and HIV literacy and varying needs of PrEP users (e.g. longer discussions about PrEP and wider sexual health issues) (12.2)  23. Facilitate and sustain an organisational culture that values a wholistic approach to sexual health and wellbeing (12.2) (e.g. reflect a wholistic approach in the sexual health service values and mission statement and include as a core competency for professional conduct, address in education sessions (5.1, 5.3, 5.6) and clinical supervision, and as part of annual appraisals (2.3, 2.4, 2.2, 2.7))  25. Establish good connections with other specialist services (e.g. delivered by NGOs, those available within the sexual health service) (12.2) that sexual healthcare professionals could signpost and/or directly refer PrEP users to, for appropriate expert support (3.1)  27. Check that PrEP users are aware of other specialist services available locally (e.g. delivered by NGOs, available within the sexual health services) and signpost or make a direct referral, as necessary (3.1) | 13. Keep but modify (discussed) – not PrEP specific but providing individualised PrEP care and responding to long standing health inequalities in health and HIV literacy and varying need is important and needs to feature somewhere (flexibility of service provision). ‘Innovative’ is too subjective. Duplicate (PA5&6 have same final recommendations so collapsed into the same row in Table 2 in paper)  23. Reject. Duplicate  25. Keep but modify (discussed) – Two-way connections / partnership work but need to word carefully so doesn’t seem like sexual health services are passing the buck and being mindful of 3^rd^ sector funding cuts. Include NGOs that serve communities other than GBMSM. Duplicate (PA5&6 have same final recommendations so collapsed into the same row in Table 2 in paper)  27. Keep but modify (discussed) – reciprocity of connection that is in stage 1. Duplicate (PA5&6 have same final recommendations so collapsed into the same row in Table 2 in paper) | PrEP services should ensure flexible provision of individualised PrEP care that meets diverse needs. For example, explore and provide ways of scheduling appointments with built-in flexibility to respond to long-standing inequalities in health and HIV/PrEP literacy during consultations  --  PrEP services and NGOs should enhance and maintain good connections across HIV prevention and care and other specialist services, to facilitate easy reciprocal referrals. Consider carefully the type of support required and which service is best placed to provide it  PrEP providers and NGO staff should support PrEP users to navigate services and online information for appropriate expert support. *Support could include signposting and/or referring PrEP users to other specialist services across and beyond the HIV prevention and care sector, as necessary* |
| -- | PrEP users find it easy to discuss wider sexual health issues because they have built a trusting relationship and familiarity with PrEP providers through continuity of care | “*R: If I go and see him, at the [clinic], he knows my situation, he's actually really good on just being able to advise. If I go to the [clinic], it's completely luck of the draw who I get. So sometimes they'll have seen me five, six years ago, and won't remember me at all. It's better having the continuity, I think.*  *I: In terms of why it's better, what does it change for you?*  *R: It just feels safer, actually, there's a bond, there's a trust going on there… I mean, you should be able to trust a doctor, but for some reason, I find actually speaking to someone that I've known for a while, actually, I feel a lot more comfortable about that*.” (PrEP user) | Environmental context and resources  Social influences  Professional role and identity | Environmental restructuring  Training  Education  Modelling  Enablement | 12.2 Restructure the social environment  12.1 Restructure the physical environment  7.1 Prompts/cues  5.1 Information about health consequences  5.3 Information about social and environmental consequences  6.1 Demonstration of the behaviour  2.2 Feedback on behaviour  2.3 Self-monitoring of behaviour | 14. Where possible, assign each PrEP user a ‘usual' sexual healthcare professional and operate a buddy system where paired sexual healthcare professionals can see each other's patients, for example, when the other is on leave, to facilitate continuity of care (12.2)  6b. Ensure the appointment system is open and the rota agreed far enough in advance to enable PrEP users to book their next PrEP appointment with their ‘usual’ sexual healthcare professional or buddy before leaving the premises (12.1)  7. Prompt sexual healthcare professionals (e.g. via paper-based or electronic checklists/ proformas, SOPs, ‘pop-up’ messages within the IT system) to remind PrEP users to book their next appointment before leaving the premises (7.1)  24. Facilitate and actively maintain (e.g. via training, huddles, clinical supervision) a warm, welcoming, and friendly atmosphere wherein sexual healthcare professionals communicate with patients in a non-judgemental manner, using inclusive, sex- and PrEP-positive, and destigmatising language to establish trust and ensure an open dialogue (12.2, 5.3)  18. Promote the advantages of high-quality clinical record keeping for continuity of care (5.1, 5.3), share best practice examples that meet the standards set out by the sexual health service and/or a relevant professional bodies (6.1), and appraise and encourage sexual healthcare professionals to reflect on their sRejects of recording episodes of care (2.2, 2.3) | 14. Reject. Duplicate  6b. Reject (not discussed) – impractical for PrEP users to have a designated sexual healthcare professional for reviews. The bit about the appointment system being open in advance is covered in 6a. Duplicate  7. Reject (discussed) – Reject all pop-ups. Already happens. Duplicate  24. Reject (discussed) – support the general sentiment but is not PrEP specific. Useful content for the intro as we will need to make a statement pointing towards existing standards / expectations of the bedrock of delivery Duplicate.  18. Reject (not discussed) – not PrEP specific. Is addressed in existing clinical governance, appraisal and revalidation. Duplicate | Not taken further  Not taken further  Not taken further  Not taken further  Not taken further  --  -- |

**Table S7 Priority area 7: BCW analysis of ‘PrEP users stay on PrEP for as long as relevant’**

| **Barriers** | **Facilitators** | **Indicative quotes** | **TDF domains** | **Intervention Functions** | **Potential BCTs**  **Numbers-=BCTs** | **Recommendations for those considering implementing PrEP at scale**  **Accept/Reject/Modify**  **Number in brackets=BCTs** | **Post APEASE and expert input decision**    **Accept/Reject/Modify** | **Agreed final recommendations for those considering implementing PrEP at scale** |
| --- | --- | --- | --- | --- | --- | --- | --- | --- |
| PrEP users find it difficult to stay on PrEP for as long as relevant because they experience or are concerned about side-effects (e.g. allergy, rash, dry mouth, GI upset, spots, longer-term renal toxicity) | -- | “*I expected those kind of symptoms with dry mouth and the wee bit funny queasiness maybe but in reality it was a lot more intense and a lot worse than what I anticipated*.” (Stopped using PrEP) | Beliefs about consequences  Behavioural regulation | Education  Enablement | 5.1 Information about health consequences  1.2 Problem solving  2.6 Biofeedback  9.2 Pros and cons  3.1 Social support | 35. Educate PrEP users (e.g. verbally at PrEP appointments, in national patient information booklets, via sexual health services, NGO, and HIV/PrEP activists’ websites and social media) about the potential side-effects of PrEP and their typically transient nature (5.1), share management strategies for the most common side-effects (1.2), and reassure against concerns about longer-term toxic effects by drawing attention to the tests undertaken at three-month reviews (5.1) and always informing them of their results (2.6)  36. Sexual healthcare professionals should discuss the various ways that PrEP users could take PrEP in an unbiased manner (5.1) and engage in a shared decision-making process to decide whether switching regimens may be appropriate (9.2)  37. Sexual healthcare professionals should inform PrEP users how to access the sexual health service for ad-hoc adherence support between appointments and ensure contact details and opening hours are kept up to date on the sexual health service website (3.1) | 35. Keep but modify (discussed) – nice wording addressing side-effect management. Again, make sure materials are co-produced and that communication routes are acceptable to key populations. Could suggest a greater proportion of discussions cover side-effects which are very real for some people and do affect adherence. Amended and merged with 36.  36. Keep but modify (discussed) – needs more focus on side-effects. Suggested wording ‘Sexual healthcare professionals should engage in well-informed and sophisticated discussions with PrEP users to create a personalised PrEP care plan”. Amended and merged with 35.  37. Keep but modify (discussed) – don’t want to encourage people to have ad-hoc appointments but people do need the confidence to navigate the healthcare system (i.e. know they can come back to speak to a sexual healthcare professional, option to change regimens). Duplicate but tweaked so final rec includes specifics re: managing side-effects and is no longer a duplicate. New rec about a list of management strategies that can be shared with PrEP users. | PrEP providers should spend an adequate proportion of PrEP discussions educating PrEP users about possible side-effects and their typically transient nature and reassure against concerns about longer-term issues and create a personalised PrEP care plan, including information on switching regimens. Reassurance can be provided by drawing attention to the regular reviews offered to PrEP users.  PrEP providers and NGO staff should support PrEP users to navigate services and online information for appropriate expert support. *Support could include: ensuring PrEP users know they can return to or call the PrEP service to discuss side-effects and have the option to change regimens; and raising awareness of and directing PrEP users to reputable online sources of side-effect management*  PrEP services should provide PrEP providers and NGO staff with a list of management strategies for common side effects that they can share with PrEP users |
| PrEP users find it difficult to stay on PrEP for as long as relevant because their sexual partner(s) is suspicious of PrEP use as they associate it with promiscuity and infidelity | -- | “*I think he thought if I was on it, I was having sex with other people, and there was that...he kind of said, well, why would you be on it if we’re together now? And in my mind, I was thinking, well, I want to be on it just to make sure*.” (Stopped using PrEP) | Social influences  SRejects | Enablement  Training | 13.2 Framing/ reframing  3.1 Social support (unspecified)  4.1 Instruction on how to perform the behaviour | 38. Ensure PrEP information and communications (e.g. national patient information booklet, posters in clinic waiting areas and consultation rooms and NGO settings, via sexual health services, NGO, and HIV/PrEP activists’ websites and social media, marketing campaign) address PrEP-related stigma, for example, by adopting ‘needs-based’ terminology rather than focusing on ‘risk’ and presenting PrEP as a responsible choice and positive means of reducing the likelihood of acquiring HIV, and include specific content on PrEP use within the context of a relationship to enable supportive and well-informed discussions among sexual partners (13.2)  30. Sexual healthcare professionals and NGO staff should encourage and support PrEP users to have wholistic conversations with their sexual partner(s) about the meaning of PrEP and the boundaries of the relationship (3.1), for instance, by sharing example phrases that they could incorporate into discussions (4.1) | 38. Keep but modify (discussed) – the first bit is not a priority here, will be addressed in stage 1. However, the last few lines are important and nice wording.  30. Keep but modify (discussed) – maintain the ethos of sexual healthcare professionals using their professional judgement to suggest/encourage and support PrEP users to have discussions with important others about PrEP and what it means in order to help people initiate (stage 2) and stay on PrEP though not necessarily something you do for everyone, tailor to the individual. Something quite novel to help support PrEP users stay on PrEP could be to recommend that sexual healthcare professionals explore and probe motivations for PrEP users wanting to stopping PrEP, including sexual partners’ reactions / views / perceptions. | PrEP information and communications should include specific content on PrEP use within the context of a relationship to address PrEP stigma, enable supportive and well-informed discussions among sexual partners, and prevent discontinuation of PrEP where there is an ongoing identified need. *Ensure that materials are co-produced and that communication routes are acceptable to key populations*  PrEP providers and NGO staff should consider sexual partners’ reactions, views, and perceptions when exploring and probing PrEP users’ motivations for wanting to stop or having stopped using PrEP, be cognisant of sexual partner influences on PrEP users’ decisions to remain on PrEP, and use their professional judgement to encourage and support PrEP users to have wholistic conversations with their sexual partner(s) about the meaning of PrEP and boundaries of the relationship(s). Share co-produced example phrases that PrEP users could incorporate into discussions |
| PrEP users find it difficult to stay on PrEP for as long as relevant because they acquire recurrent STIs while on PrEP | -- | “*PrEP coming around allowed them [patients] to feel that they could have more sex, with different people, and not use condoms, and not have to sit and panic about it either. And some of them have come back subsequently and said, oh I'm getting other infections, I don't like this anymore, I'm going back to using condoms*.” (Sexual healthcare professional) | Beliefs about consequences | Education  Persuasion | 5.1 Information about health consequences  13.2 Framing/ reframing | 28. Sexual healthcare professionals and NGO staff should advise PrEP users that PrEP only protects against HIV and present the likelihood of contracting an STI following non-condom protected intercourse, taking account of various risk factors (5.1)  29. Sexual healthcare professionals and NGO staff should encourage PrEP users to continue using condoms alongside PrEP by framing PrEP as an additional rather than alternative HIV prevention method (13.2) | 28. Reject (discussed) – already in national guidelines.  29. Reject (discussed) – there is some dissonance / a PrEP paradox here as the majority of users are eligible because of condomless sex… so if they start using condoms then they may no longer be eligible. Condoms should be mentioned as part of combination prevention or, for the majority of PrEP users, as an option for STI prevention. But framing it as additional HIV prevention could undermine the message about the effectiveness of PrEP. | Not taken forwards  Not taken forwards  -- |
|  | PrEP users find it easy to stay on PrEP for as long as relevant because of the positive health, emotional, and social consequences of PrEP (e.g. effectively safeguards their own and other people’s sexual health, reassures against personal HIV fear, social acceptability) | “*I don’t see that [stopping PrEP] being something I would consider in the short to medium term. I just feel that it gives me reassurance, both in terms of medical reassurance but also psychological reassurance*.” (PrEP user) | Beliefs about consequences | Education  Persuasion | 5.1 Information about health consequences  5.3 Information about social and environmental consequences  5.6 Information about emotional consequences | 02. PrEP information and communications (e.g. national patient information booklet, posters in clinic waiting areas and consultation rooms and NGO settings, via sexual health services, NGO, and HIV/PrEP activists’ websites and social media, marketing campaign) should include education on the positive health impacts of PrEP, as well as the wider social and emotional benefits and value of PrEP, for communities and individuals (5.1, 5.3, 5.6). | 02. PrEP information and communications (e.g. national patient information booklet, posters in clinic waiting areas and consultation rooms and NGO settings, via sexual health services, NGO, and HIV/PrEP activists’ websites and social media, marketing campaign) should include education on the positive health impacts of PrEP, as well as the wider social and emotional benefits and value of PrEP, for communities and individuals (5.1, 5.3, 5.6). | PrEP information and communications should include education on the positive health impacts of PrEP, as well as the wider social and emotional benefits and value of PrEP, for communities and individuals |

### Table S8 Priority area 8- A BCW analysis of ‘PrEP providers communicate the decision to not provide further PrEP’

| **Barriers** | **Facilitators** | **Indicative quotes** | **TDF domains** | **Intervention Functions** | **Potential BCTs**  **Numbers = BCTs** | **Recommendations for those considering implementing PrEP at scale**  **Accept/Reject/Modify**  **Number in Brackets = BCTs** | **Post APEASE and expert input decision**    **Accept/Reject/Modify** | **Agreed final recommendations for those considering implementing PrEP at scale** |
| --- | --- | --- | --- | --- | --- | --- | --- | --- |
| PrEP providers find it difficult to communicate the decision to not provide further PrEP because of inadequate discussion with PrEP users about the risk-benefit of PrEP at the outset, owing to a lack of knowledge, sRejects, and experience | PrEP providers find it easy to communicate the decision to not provide further PrEP because they mention at the start that need for PrEP may change over time (i.e. circumstance-dependent) and that ongoing eligibility will be assessed and is required to keep issuing PrEP | **Example 1**  “*It becomes an issue when there are some reasons maybe not to give PrEP, there are some side-effects, or there's some effect on renal function. And then having to go back and talk about the risk-benefits again. In lots of people, that tends to be not fully discussed properly, it’s kind of glossed over*.” (Sexual healthcare professional)  **Example 2**  “*We advise that there will be continuing assessments of their eligibility and it may be that they will drop in and out of eligibility depending on their sexual risk because we understand that that changes with time for everybody. So we do make it very clear that that will happen*.” (Sexual healthcare professional) | Knowledge  SRejects  Professional role and identity | Education  Training  Persuasion  Modelling | 5.1 Information about health consequences  4.1 Instructions on how to perform the behaviour  7.1 Prompts/cues  6.1 Demonstration of the behaviour  8.1 Behavioural practice/rehearsal  2.2 Feedback on behaviour  5.3 Information about social and environmental consequences  9.1 Credible source | 20. Ensure sexual healthcare professionals are educated, trained, and appraised in their sRejects in discussing the risks and benefits of PrEP (e.g. through online modules, peer support, clinical supervision), for example, by giving information on PrEP health consequences (5.1), producing a ‘how to’ script for common PrEP scenarios based on the lessons learned of sexual healthcare professionals with general medicine expertise (4.1, 7.1), and providing opportunities to shadow (6.1), practice with (8.1), and receive feedback (2.2) from more experienced sexual healthcare professionals  15. Mandate discussion with PrEP users about the risk-benefit of PrEP in a formal protocol that details the key activities required to be completed by sexual healthcare professionals at the initial PrEP assessment (4.1, 5.1)  21. Share positive testimonials (e.g. via emails, intranet) of respected senior sexual healthcare professionals promoting the social benefits of communicating the risk-benefit of PrEP, idea of ‘seasons of risk’, and requirement for checks on and ongoing need for PrEP at the initial PrEP consultation (5.3, 9.1) | 20. Keep but modify (discussed) – too generic in its current form, applies to all clinical care. Tighten to be more specific to PrEP e.g. training for sexual healthcare professionals to understand and explain instances in which stopping PrEP may be in the PrEP user’s best interests (think about the wording) and a UKMEC style PrEP document with examples of clear situations where the risk outweighs the benefits that sexual healthcare professionals can refer to. *  15. Reject (discussed) – already part of clinical governance/ competence. Worry that it separates PrEP from broader combination prevention. More important recommendations to prioritise.  21. Reject (not discussed) – part of clinical governance / CPD. | PrEP services should use multi-methods to develop PrEP providers’ knowledge of and Rejects in explaining instances when stopping PrEP may be in a PrEP user’s best interests. *For example, develop and educate PrEP providers on guidance that includes examples of situations where the risk of PrEP outweighs the benefits, co-produce scripts that address a range of literacy needs for common PrEP risk-benefit scenarios, and provide opportunities to shadow, practice, and receive feedback on communicating decisions to stop PrEP*  --  -- |

### Table S9 - Priority area 9- BCW analysis of ‘PrEP providers explore PrEP users’ reasons for wanting to stop / stopping using PrEP’

| **Barriers** | **Facilitators** | **Indicative quotes** | **TDF domains** | **Intervention Functions** | **Potential BCTs**  **Numbers = BCTs** | **Recommendations for those considering implementing PrEP at scale**  **Accept/Reject/Modify**  **Numbers in brackets =BCTs** | **Post APEASE and expert input decision**    **Accept/Reject/Modify** | **Agreed final recommendations for those considering implementing PrEP at scale** |
| --- | --- | --- | --- | --- | --- | --- | --- | --- |
| PrEP providers find it difficult to explore PrEP users’ reasons for wanting to stop / stopping using PrEP because PrEP users tend not to return and discuss their decision | PrEP providers find it easy to explore PrEP users’ reasons for wanting to stop / stopping using PrEP because there are follow-up and/or other targeted intervention processes in place | “*Generally, we wouldn’t see them again, they just don't access the service, because obviously they feel they don't need it at the moment. So, they don't need PrEP, and they’ve not been for a sexual health screen. But if they do come back for a sexual health screen, then we'd say, I see you’ve dropped your PrEP, why was that. And kind of just reflect on it with them, is that the decision that they're happy with, and do they still want to remain off PrEP*.” (Sexual healthcare professional) | Behavioural regulation  Environmental context and resources | Education  Enablement  Environmental restructuring | 5.1 Information about health consequences  9.1 Credible source  1.8 Behavioural contract  1.9 Commitment  1.1 Goal setting (behaviour)  7.1 Prompts/cues  12.2 Restructure the social environment | 31b. Use a multi-method approach, including posters, national patient information booklets, positive testimonials of PrEP users, online resources, and verbal communication by sexual healthcare professionals and NGO staff, to emphasise the importance of PrEP users discussing stopping PrEP with a sexual healthcare professional (5.1, 9.1)  01. Sexual healthcare professionals could ask PrEP users to verbally agree to or sign a written contract specifying that they will attend for regular PrEP reviews, even if they have stopped PrEP in the interim period (1.8, 1.9, 1.1)  9. Integrate ‘pop-up’ messages into the IT system to alert sexual healthcare professionals that the patient presenting at their clinic did not attend or is overdue a PrEP appointment and advise them to raise the issue and explore if they have stopped using PrEP and if so, why (7.1)  10b. Run a monthly report on the IT system to identify ‘did not attends’ and those overdue a PrEP appointment and attempt to make contact (e.g. via email, phone, SMS) and discuss their decision to stop using PrEP, if applicable (12.2) | 31b. Reject (not discussed) –emphasising the importance of discussing stopping PrEP with a sexual healthcare professional is already included as part of 31a. No point repeating so makes sense to include for PA3 ‘PrEP users attend PrEP reviews’ rather than for this PA in Table 2.  01 -REJECT HP only - impractical and too paternalistic  9. Reject (discussed) – favour automated system which sends reminders to PrEP users. Duplicate.  10b. Keep (discussed) – about knowing your PrEP cohort (at service-level). | Not taken further  --  --  PrEP services should assess monitoring and evaluation data to identify ‘did not attends’ and those overdue a PrEP review and attempt to make contact to discuss decisions to stop using PrEP and reengage them with PrEP care, as appropriate |

### Table S10 - Priority area 10- A BCW analysis of ‘PrEP users stop using PrEP’

| **Barriers** | **Facilitators** | **Indicative quotes** | **TDF domains** | **Intervention Functions** | **Potential BCTs**  **Numbers = BCTs** | **Recommendations for those considering implementing PrEP at scale**  **Accept/Reject/Modify**  **Numbers in brackets =BCTs** | **Post APEASE and expert input decision**  **Accept/Reject/Modify** | **Agreed final recommendations for those considering implementing PrEP at scale** |
| --- | --- | --- | --- | --- | --- | --- | --- | --- |
| PrEP users find it difficult to stop using PrEP because of the social acceptability of PrEP and emerging stigmas around *not* using PrEP | -- | “*The decision to come off [PrEP] is much harder and more layered than deciding to go on it in the first place…I’ve got an option to continually be safe and have that faith in it, why would I ditch that faith in it. So there’s guilt there personally. But, again with Grindr…it’s a bit like, well if I’m changing my setting to [HIV] negative instead of being on PrEP, what am I saying? Am I basically saying, one that I’m not valuing my own sexual health and two am I not valuing their sexual health*?” (Stopped using PrEP) | Beliefs about consequences | Education  Persuasion | 5.1 Information about health consequences  13.2 Framing/ reframing | 33. Use a range of educational methods to enhance PrEP users’ understanding of behaviours and situations that carry a higher likelihood of acquiring HIV and facilitate accurate assessments of when they no longer have a need for PrEP (5.1)  34. PrEP information and communications (e.g. interactions with sexual healthcare professionals and NGO staff, national patient information booklets, sexual health services, NGO, and HIV/PrEP activists’ websites and social media, marketing campaigns) should address emerging stigmas around people *not* using PrEP by framing it as an additional rather than alternative HIV prevention method (i.e. one of many options) (13.2) and sharing information on the effectiveness of alternative sexual health promotion methods that offer sufficient protection against HIV (5.1) | 33. Reject (discussed) – more important to encourage people to start PrEP than to stop. It’s also incredibly difficult to assess risk for non-GBMSM PrEP users. Duplicate.  34. Keep but modify (discussed) – the focus should be on the importance of offering choices and explaining the ‘seasons of risk’ concept because of emerging stigmas around *not* using PrEP. Inform people of all options for HIV prevention and ensure information and communications are tailored to the needs of distinct key populations (bearing in mind many of those who stand to benefit from PrEP may not be reached by current channels). | Not taken further  PrEP and wider sexual health resources and communications should inform of all options for HIV prevention, emphasise the importance of choices, and explain the ‘seasons of risk’ concept to address emerging stigmas around *not* using PrEP. *Ensure that materials are co-produced and that communication routes are acceptable to key populations* |
| -- | PrEP users find it easy to stop using PrEP because of a change in their self-perceived HIV risk (e.g. not planning any sexual activity, in a monogamous relationship) | “*We just got to the point in the relationship where we had a discussion about being exclusive, about sex, about safe sex and made a decision not to see anybody else, be monogamous, and I then took the decision to come off PrEP because I didn’t think I needed it anymore*.” (Stopped using PrEP) | Beliefs about consequences | Education  Persuasion | 5.1 Information about health consequences | 33. Use a range of educational methods to enhance PrEP users’ HIV literacy and ensure they have an accurate understanding of behaviours and situations that carry a higher likelihood of acquiring HIV (5.1) | 33. Reject (discussed) – more important to encourage people to start PrEP than to stop. It’s also incredibly difficult to assess risk for non-GBMSM PrEP users. Duplicate. | Not taken further |
